# Single-cell profiling of human plaques reveals cellular and molecular signatures of symptomatic atherosclerosis

**DOI:** 10.64898/2026.11.07.26367408

**Authors:** Anushree Ray, Luka Živković, Lanyue Zhang, Roya Batool, Abdalla Marei, Martin Dichgans, Nikolaos Tsilimparis, Marios K. Georgakis

## Abstract

Atherosclerotic plaque vulnerability drives ischemic cardiovascular events, yet the cellular and molecular signatures distinguishing symptomatic from asymptomatic human plaques remain poorly defined. Here, we compared 53 symptomatic and asymptomatic human carotid plaques using single-nucleus (snRNA-seq; 17 samples, 19,100 nuclei; AtherOMICS biobank) and single-cell RNA sequencing (scRNA-seq; 36 samples, 68,983 cells; four public datasets), analysed independently and combined by meta-analysis. Across sequencing approaches, symptomatic plaques showed cell-state shifts toward later or terminal states in osteochondrogenic smooth muscle cells (SMCs) and CD4⁺ T cells, as well as transcriptional changes toward inflammatory and epithelial-to-mesenchymal transition (EMT) in endothelial cells (ECs), T cells, and fibroblasts. Nuclear profiling uncovered a depletion of fibroblasts in symptomatic plaques, marked by downregulation of *RGS5*. Integrating these transcriptomic signatures with genome-wide association data for coronary, cerebrovascular, and peripheral atherosclerotic disease, we found that genetic risk was enriched primarily in vascular wall cells (SMCs, fibroblasts, ECs), rather than immune cell types. The analyses further nominated higher *TWIST1* expression in ECs (colocalized with risk across four atherosclerotic phenotypes), lower *GUCY1A1* expression in fibroblasts and macrophages, and higher *MRAS* expression in T cells as candidate driver mechanisms linking genetic risk to symptomatic plaque transition. These findings provide novel insights into cellular and molecular phenotypes associated with plaque vulnerability and identify potential driver mechanisms of symptomatic plaque transformation that could inform therapeutic development.

## INTRODUCTION

Atherosclerosis is the principal pathology underlying ischemic cardiovascular diseases, which remain the leading causes of death and disability worldwide.^1,2^ Atherosclerotic plaques arise within the subendothelial space and exhibit marked heterogeneity in clinical behavior.^3^ Plaque vulnerability is thought to be a major driver of this heterogeneity,^4^ yet the mechanisms governing plaque fate remain incompletely understood and are not directly targeted by current therapies. Histological hallmarks of plaque vulnerability do not fully explain divergent clinical trajectories, motivating the search for cellular and microenvironmental determinants that may drive plaque destabilization.^5^

Single-cell transcriptomic studies have provided novel insights into the cellular and molecular architecture of human atherosclerosis, revealing greater cellular and functional diversity than is captured by histology or bulk profiling.^6–8^ For example, they have revealed that smooth muscle cells (SMCs) undergo phenotypic transition toward fibromyocyte and osteochondrogenic states.^9,10^ Macrophages resolve into inflammatory, lipid-associated foamy, heme-handling, and remodeling populations with distinct functional programs,^11^ whereas endothelial cells (ECs) stratify into quiescent, proangiogenic, and endothelial-to-mesenchymal transition (EndoMT)-associated states linked to vascular remodeling and neovascularization.^6,7^ Despite these advances, existing studies have largely been limited by sample size and have not systematically associated cellular and molecular features with clinical trajectories. Consequently, the cell states and microenvironmental programs associated with plaque progression and destabilization remain incompletely defined.

Here, we compared symptomatic and asymptomatic human carotid plaques from 53 samples using single-nucleus (snRNA-seq; n = 17; AtherOMICS biobank^12^) and single-cell RNA sequencing (scRNA-seq; n = 36; four public datasets), analysing each modality separately and meta-analysing comparable estimates. We systematically compared cell-type composition, transcriptional programs, pathway activity, and lineage trajectories across modalities, clinical phenotypes, and established histological features of plaque vulnerability. By integrating our results with data from genome-wide association studies (GWAS), we further detected cell-specific differentially expressed genes (DEGs) in symptomatic plaques that might drive genetic risk for atherosclerotic cardiovascular disease.

## RESULTS

### Data resources and study design

We leveraged in-house snRNA-seq data and published scRNA-seq from human carotid plaques to characterize features of cellular organization and gene expression, and then investigated associations between these features and symptomatic plaque status (**Fig. 1**). In the AtherOMICS biobank, we performed snRNA-seq on carotid plaque samples from 17 patients undergoing endarterectomy.^12^ Eight samples were classified as symptomatic based on a history of ipsilateral ischemic stroke, transient ischemic attack, central retinal artery occlusion, or amaurosis fugax. **Supplementary Table 1** presents the demographic and clinical characteristics of AtherOMICS patients. After quality control, doublet removal, and variance-stabilized normalization, 19,100 nuclei were retained for downstream analyses. In parallel, we identified four publicly available carotid plaque datasets that included sufficient individual-level information to classify samples as symptomatic or asymptomatic. These datasets comprised 36 carotid plaque samples, of which 16 were classified as symptomatic. After harmonized preprocessing and quality control, 68,983 cells were retained. Using a unified annotation and analytical framework, we compared symptomatic and asymptomatic plaques across multiple levels, including cell-type composition, cell-state-specific gene expression, pathway activity, and lineage progression.

**Fig. 1.**
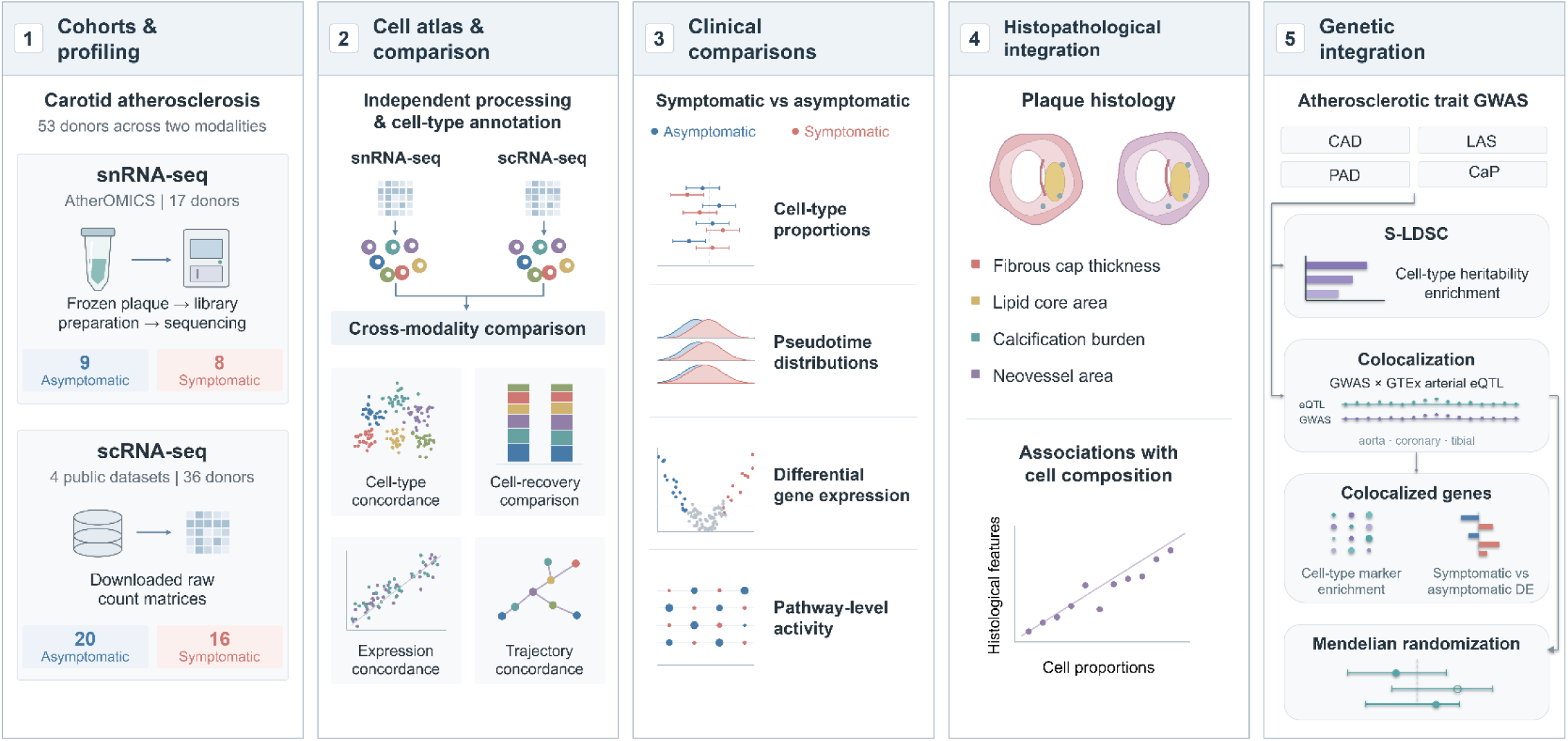
Study design. (1) Cohorts and profiling. Carotid atherosclerosis was studied in 53 samples across two modalities. snRNA-seq (AtherOMICS): 17 carotid endarterectomy samples (9 asymptomatic, 8 symptomatic); nuclei isolation from frozen plaque, library preparation and sequencing. scRNA-seq: 36 samples from four public datasets (20 asymptomatic, 16 symptomatic); raw count matrices retrieved from public repositories. (2) Cell atlas and comparison. Each modality was quality-controlled, clustered and annotated independently. This was followed by four cross-modality comparisons: cell-type concordance, cell-type recovery, pseudobulk expression concordance and lineage trajectory concordance. These comparisons provided the modality-specific reference used to interpret downstream symptomatic-versus-asymptomatic analyses. (3) Clinical comparisons. Symptomatic and asymptomatic plaques were compared independently in each modality, with meta-analysis where applicable. The comparisons covered cell-type proportions, pseudotime distributions and terminal-state shifts, pseudobulk differential expression and Hallmark pathway enrichment. (4) Histological integration. snRNA-seq cell-type proportions were correlated with quantitative histological features: fibrous cap thickness, lipid-core area, calcification burden and neovessel area. (5) Genetic integration. GWAS summary statistics for CAD, LAS, PAD and CaP were analysed using stratified linkage disequilibrium score regression (S-LDSC) for cell-type-specific heritability enrichment, and colocalization with GTEx v8 arterial eQTL data (aorta, coronary and tibial artery) across all four traits (12 trait–artery combinations). Colocalized genes were then tested for cell-type marker enrichment and examined for cell-type-specific differential expression between symptomatic and asymptomatic plaques. scRNA-seq, single-cell RNA sequencing; snRNA-seq, single-nucleus RNA sequencing; GWAS, genome-wide association study; CAD, coronary artery disease; LAS, large artery stroke; PAD, peripheral artery disease; CaP, Carotid plaque presence; S-LDSC, stratified linkage disequilibrium score regression; eQTL, expression quantitative trait locus

### Cellular landscape of human carotid plaques in single-cell and single-nucleus RNA sequencing

After harmonized preprocessing and quality control, we assigned broad and fine cell-type annotations using canonical marker genes derived from published single-cell studies and atlases of human vascular tissue and atherosclerotic plaques.^9,11,13–18^ Uniform manifold Approximation and Projection (UMAP) embeddings identified seven broad plaque cell types that were consistently detected across both scRNA-seq and snRNA-seq datasets: SMCs, ECs, fibroblasts, macrophages, T cells, mast cells, and plasma cells (**Fig. 2a**). Small populations of natural killer cells and B cells were detected only in the scRNA-seq datasets and were not recovered by snRNA-seq. At the subtype level, annotations revealed a largely conserved cellular state architecture across modalities (**Supplementary Fig. 1**). SMC states comprised contractile SMCs, fibromyocyte SMCs (fibroblast-like, cap-stabilizing), and osteochondrogenic SMCs (phenotypically modulated, microcalcification-associated). EC states included quiescent ECs, proangiogenic ECs (neovessel-forming, intraplaque angiogenesis-associated), and EndoMT ECs (mesenchymal-transitioning). Macrophage states comprised homeostatic macrophages (tissue-resident, non-activated), TREM2⁺ foamy macrophages (lipid-associated), PLIN2⁺/TREM1⁺ macrophages (inflammatory, lipid-associated), HMOX1⁺ heme-handling macrophages, C1Q⁺ remodeling macrophages (efferocytic), and IL1B⁺ inflammatory macrophages (classically activated, cytokine-secreting), with IL1B⁺ inflammatory macrophages detected exclusively in scRNA-seq. Finally, T-cell states spanned naïve T cells, CD4⁺ T cells, and CD8⁺ T cells, with scRNA-seq providing further resolution into CD4⁺ effector and regulatory (Tregs) states, and CD8⁺ cytotoxic states.

**Fig. 2.**
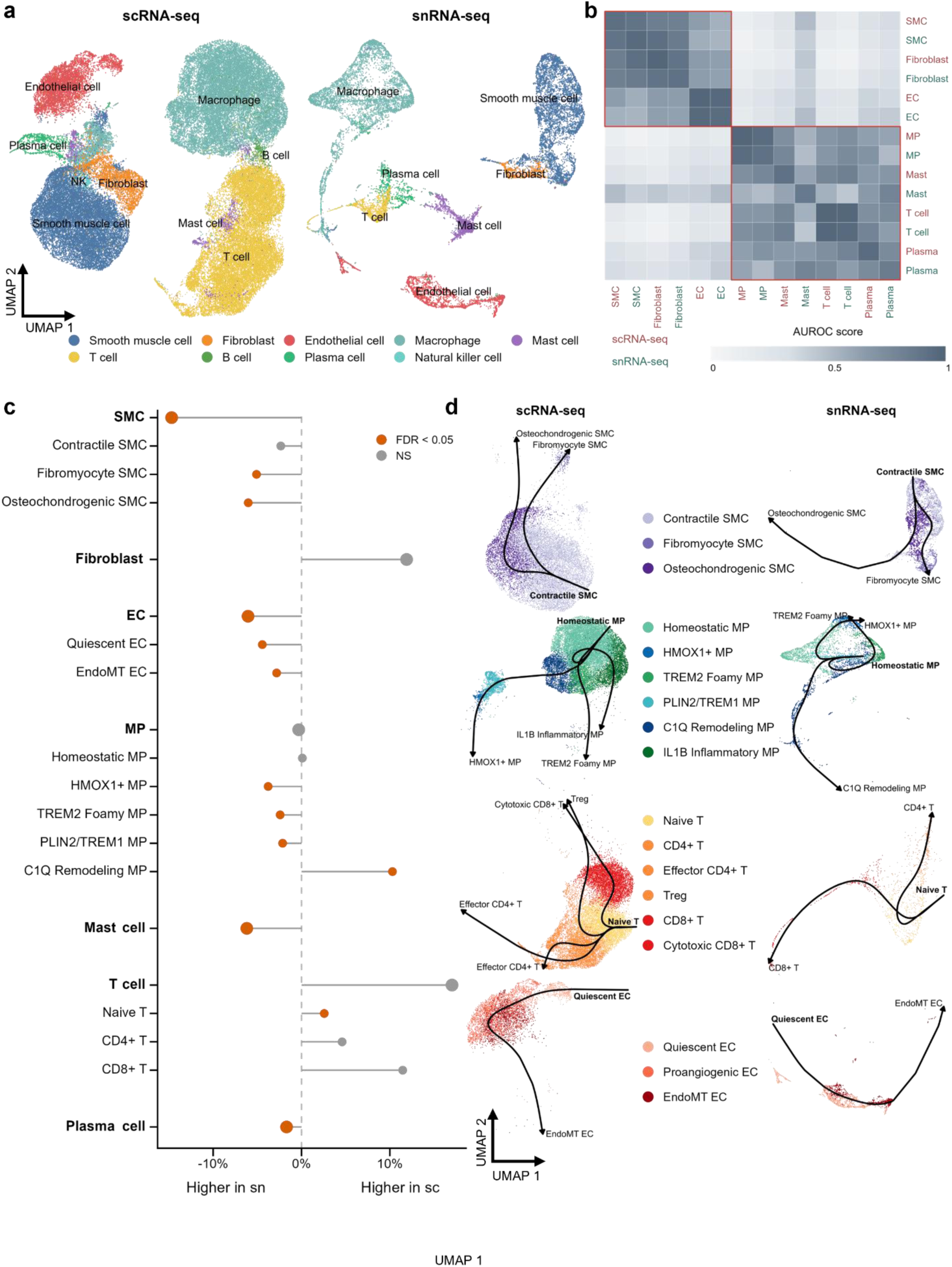
scRNA-seq and snRNA-seq recover shared plaque lineages with modality-dependent cell-type representation. (a) UMAP embeddings of broad cell-type annotations in scRNA-seq and snRNA-seq carotid plaque datasets. Cells or nuclei are coloured by broad lineage identity. (b) Cross-modality cell-type discriminability analysis using MetaNeighbor. The heatmap shows AUROC scores for pairwise cell-type similarity within and between scRNA-seq and snRNA-seq. Rows and columns are ordered by hierarchical clustering. Higher AUROC values indicate greater transcriptional similarity. (c) Difference in mean cell-type proportion between scRNA-seq and snRNA-seq across broad and fine cell types. Lollipop segments extend from zero to the estimated mean difference. Orange points indicate FDR < 0.05; grey points indicate non-significant comparisons. Positive values indicate higher recovery in scRNA-seq and negative values indicate higher recovery in snRNA-seq. (d) Slingshot trajectories inferred separately in scRNA-seq and snRNA-seq for SMCs, MPs, T cells and ECs. Cells or nuclei are shown on lineage-specific UMAP embeddings and coloured by fine cell-state annotation. Black curves indicate inferred lineage trajectories rooted at predefined biological starting states. UMAP, uniform manifold approximation and projection; scRNA-seq, single-cell RNA sequencing; snRNA-seq, single-nucleus RNA sequencing; SMC, smooth muscle cell; EC, endothelial cell; MP, macrophage; NK, natural killer; EndoMT, endothelial-to-mesenchymal transition; C1Q, complement component 1q; Treg, regulatory T cell; AUROC, area under the receiver operating characteristic curve; FDR, false discovery rate; NS, not significant

To assess the concordance of lineage annotations across modalities, we quantified pairwise cell-type discriminability using area under the receiver operating characteristic curve (AUROC) scores derived from single-cell and single-nucleus expression profiles (**Fig. 2b**). Within each modality, self-discrimination was high across annotated cell types (scRNA-seq: 0.923–0.994; snRNA-seq: 0.851–0.960), indicating strong transcriptional separation between lineages. Cross-modal AUROC scores for matching cell types were similarly high for macrophages (0.954), T cells (0.975), ECs (0.967), SMCs (0.911), and fibroblasts (0.908), with within-to-cross modality differences of ≤0.08. These findings support strong transcriptional concordance of the broad plaque lineages between scRNA-seq and snRNA-seq modalities. In contrast, mast cells (0.619) and plasma cells (0.763) exhibited lower cross-modal concordance, with within-to-cross AUROC reductions of 0.30 and 0.19, respectively, suggesting greater modality sensitivity among these less abundant immune populations. Hierarchical clustering of the complete AUROC matrix further separated vascular wall cell populations (SMCs, fibroblasts, and ECs) from immune populations (macrophages, T cells, mast cells, and plasma cells), with cross-compartment AUROC scores generally ≤0.5 across modalities. Collectively, these results demonstrate that snRNA-seq and scRNA-seq capture a highly concordant cellular landscape of human carotid atherosclerotic plaques and support robust cross-platform integration of broad plaque cell lineages.

We next examined whether scRNA-seq and snRNA-seq differed in their ability to recover plaque cell populations (**Fig. 2c**). SnRNA-seq recovered more vascular wall populations, most markedly SMCs (30.4% versus 15.6% in scRNA-seq), including fibromyocyte (5.4% versus 0.3%) and osteochondrogenic states, as well as ECs. Total macrophage proportions were similar (32.0% versus 31.7%), but subtype composition differed: lipid-associated and heme-handling states were enriched in snRNA-seq, whereas C1Q⁺ remodelling macrophages were enriched in scRNA-seq. Percentages denote mean proportions of all analyzed cells or nuclei assigned to the relevant annotation set.

To determine whether both modalities captured comparable transcriptional state transitions within broad plaque lineages, we performed lineage-specific pseudotime analyses using Slingshot (**Fig. 2d**). For each lineage, trajectories were rooted in biologically defined progenitor states, including contractile SMCs, homeostatic macrophages, naïve T cells, and quiescent ECs. SMC trajectories in both modalities defined a continuum extending from contractile SMCs toward fibromyocyte and osteochondrogenic states. Macrophage trajectories similarly connected homeostatic, inflammatory, and remodeling-associated states, although their topology differed between modalities. scRNA-seq resolved distinct branches toward IL1B⁺ inflammatory, TREM2⁺ foamy, and HMOX1⁺ heme-handling macrophage states, whereas snRNA-seq emphasized a continuum linking homeostatic and foamy macrophages with extensions toward HMOX1⁺ heme-handling and C1Q⁺ remodeling populations. T-cell trajectories spanned naïve, CD4⁺, and CD8⁺ states, connecting early and terminal differentiation states, with scRNA-seq further resolving CD4⁺ effector, Treg, and CD8⁺ cytotoxic terminal states. ECs formed a continuous trajectory from quiescent to EndoMT ECs in both modalities. These analyses indicate that broad cell-type populations are organized along comparable, continuous phenotypic trajectories in both modalities, although the branching complexity of these trajectories differs between scRNA-seq and snRNA-seq.

### Single-cell and single-nucleus RNA sequencing capture distinct transcriptional programs

We next characterized modality-associated differences that could influence interpretation of subsequent symptom-associated transcriptional analyses. Across all shared genes, mean normalized expression showed only modest concordance between scRNA-seq and snRNA-seq at the global level (R² = 0.24; **Fig. 3a, Supplementary Fig. 2**), with several inflammatory and metabolic pathways showing greater representation in scRNA-seq (**Fig. 3b, Supplementary Table 2**). Across all cell types, genes upregulated in scRNA-seq were consistently enriched for cytosolic annotations (Odds ratio [OR] = 4.35–90.75; all false discovery rate (FDR) ≤ 6.81 × 10⁻⁹), whereas genes upregulated in snRNA-seq were enriched for nuclear annotations (OR = 3.46–26.83; all FDR ≤ 0.0499; **Fig. 3c**). scRNA-seq exhibited higher overall (Unique Molecular Identifier) UMI counts and gene detection per barcode (**Fig. 3d**), highlighting differences in transcript recovery. On the other hand, dissociation stress-associated transcriptional signatures, which reflect immediate-early and cytosolic activation programs induced during enzymatic tissue dissociation rather than native *in situ* biology, were systematically elevated in scRNA-seq data across all cell types (**Fig. 3e**). Mean log_2_ fold changes (FCs) between scRNA-seq and snRNA-seq revealed larger shifts in immune cell-type populations than in vascular wall cell-type populations. Effect sizes were largest in mast cells (mean log₂FC = 4.36, 95% confidence interval [CI] = 3.87–4.86) and T cells (4.11, 3.49– 4.73), and smallest in ECs (2.09, 1.69–2.49). These findings indicate that transcriptomic differences between scRNA-seq and snRNA-seq arise from both biological compartment representation and processing-induced activation, implying the importance of modality-aware interpretation in downstream analyses.

**Fig. 3.**
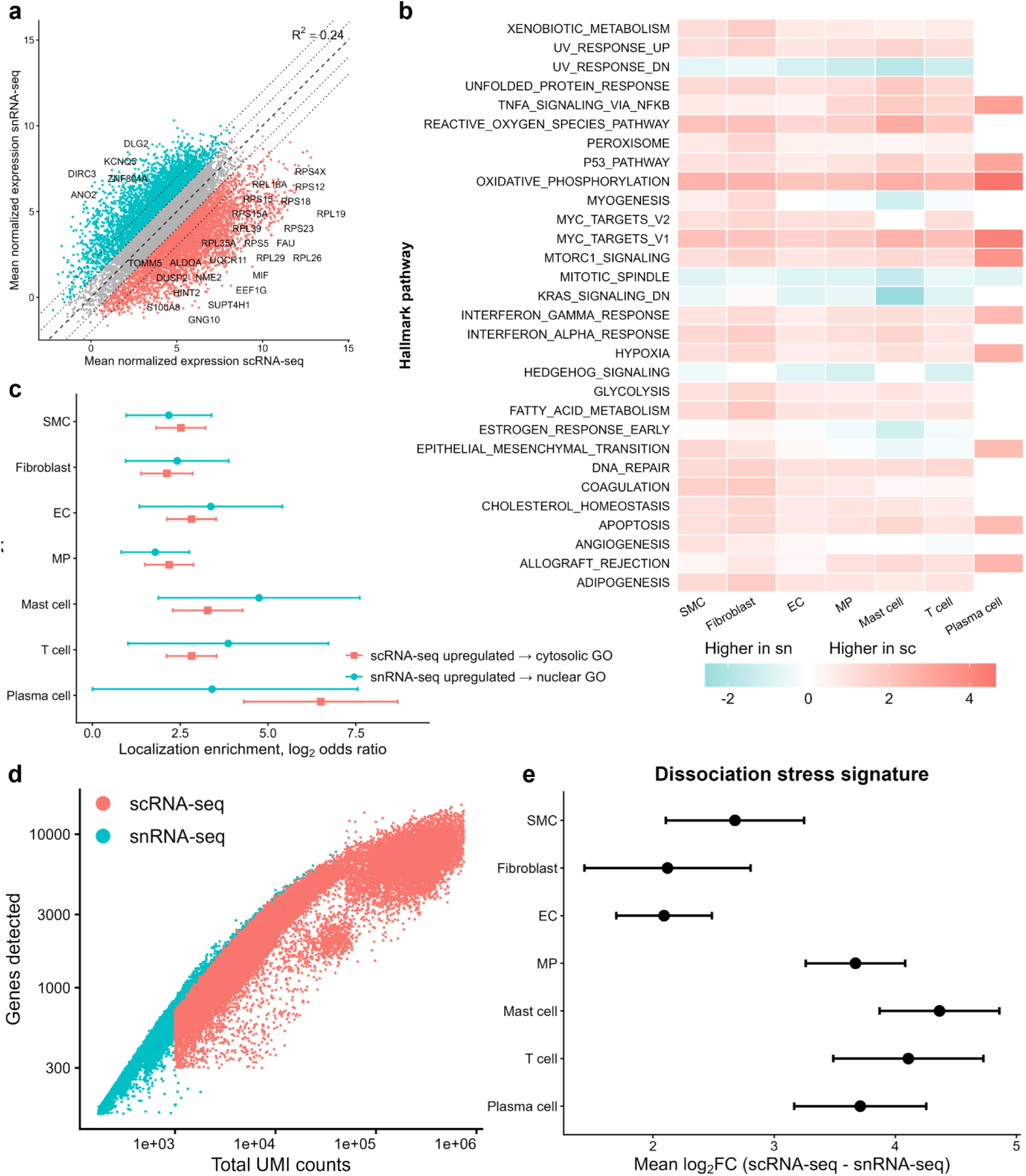
Whole-cell and nuclear profiling capture distinct transcriptional programs. (a) Gene-level concordance of mean normalized expression between scRNA-seq and snRNA-seq across shared genes. Genes are coloured by relative enrichment in scRNA-seq or snRNA-seq; selected modality-biased genes are labelled. Dotted lines indicate fold-change intervals around the dashed identity line. R² denotes the squared Pearson correlation coefficient. (b) Hallmark pathway-level (MSigDB collection H) modality effects across broad cell types. Heat map shows mean log₂FC for scRNA-seq relative to snRNA-seq across genes assigned to the 30 Hallmark gene sets with the largest absolute mean log₂FC. (c) Enrichment of subcellular localization annotations among genes upregulated in scRNA-seq or snRNA-seq. Points indicate Fisher enrichment log₂ odds ratios and horizontal bars indicate 95% confidence intervals. (d) Number of detected genes as a function of total UMI count in scRNA-seq and snRNA-seq barcodes. (e) Dissociation stress-signature differences between scRNA-seq and snRNA-seq across broad cell types. Points indicate the mean stress-gene log₂ fold change for scRNA-seq relative to snRNA-seq; horizontal bars indicate 95% confidence intervals. Positive values indicate higher stress-associated expression in scRNA-seq. scRNA-seq, single-cell RNA sequencing; snRNA-seq, single-nucleus RNA sequencing; SMC, smooth muscle cell; EC, endothelial cell; MP, macrophage; GO, Gene Ontology; FC, fold change; UMI, unique molecular identifier; R², coefficient of determination; UV, ultraviolet; TNFα, tumor necrosis factor alpha; NF-κB, nuclear factor kappa B; IFNα, interferon alpha; IFNγ, interferon gamma

### Symptomatic plaques are characterized by fibroblast depletion and trajectory shifts towards CD4⁺ T-cell, osteochondrogenic SMC and EndoMT states

We next compared cellular compositions between symptomatic and asymptomatic plaques across the scRNA-seq and snRNA-seq datasets (**Fig. 4a**). Fibroblasts were less abundant in symptomatic plaques in snRNA-seq (OR = 0.73, 95% CI = 0.56–0.94), and in the meta-analysis with scRNA-seq data (OR = 0.75, 95% CI = 0.59–0.96, P = 0.020, heterogeneity index (I²) = 0%). This association did not parallel the baseline modality-recovery pattern, as fibroblasts were not preferentially recovered by snRNA-seq, and the cross-modality estimate showed no heterogeneity. No other broad or fine cell populations showed significant differences in abundance between symptomatic and asymptomatic plaques.

**Fig. 4.**
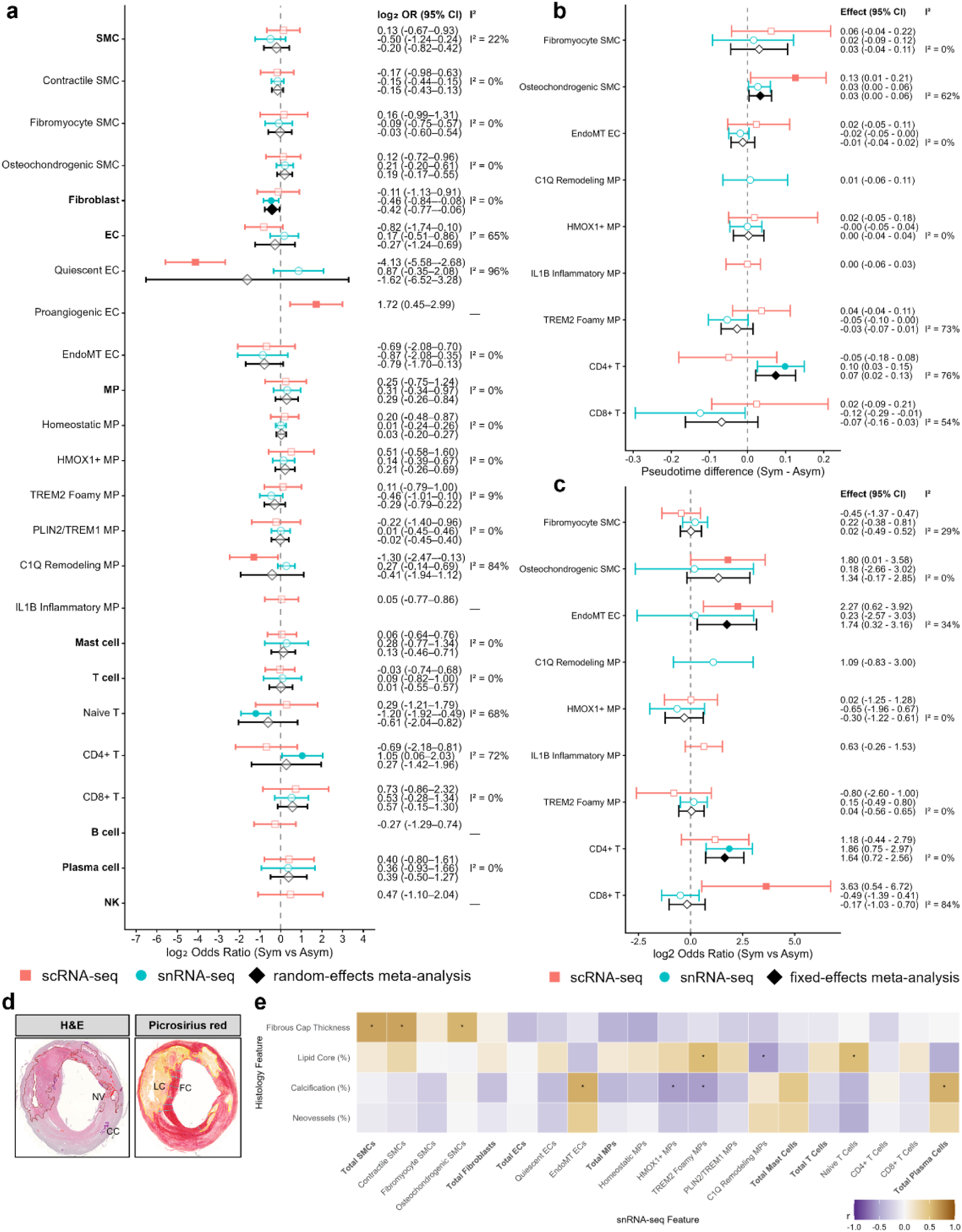
Composition and pseudotime shifts toward osteochondrogenic SMC and CD4⁺ T-cell states in symptomatic plaques correlate with histological features of plaque vulnerability. (a) Forest plot of compositional differences between symptomatic and asymptomatic plaques across broad and fine cell-type annotations. Points show log₂ OR from beta-binomial models in scRNA-seq (orange) and snRNA-seq (teal), and random-effects meta-analysis estimates (black diamonds); horizontal bars indicate 95% CIs. Filled points indicate FDR < 0.05. I² denotes between-modality heterogeneity for meta-analyzed estimates. (b) Forest plot of sample-level weighted median pseudotime differences (symptomatic − asymptomatic) across SMC, EC, MP, and T-cell lineage branches. Points show median pseudotime difference in scRNA-seq (orange) and snRNA-seq (teal), and fixed-effects meta-analysis estimates (black diamonds); horizontal bars indicate 95% CIs. (c) Forest plot of sample-level terminal-state fraction differences (symptomatic − asymptomatic) across SMC, EC, MP, and T-cell lineage branches. Points show log₂ ORs in scRNA-seq (orange) and snRNA-seq (teal), and fixed-effects meta-analysis estimates (black diamonds); horizontal bars indicate 95% CIs. (d) Representative H&E and Picrosirius red staining of a carotid endarterectomy plaque, annotated for neovessels (NV), calcification (CC), lipid core (LC), and fibrous cap (FC). (e) Pearson correlations between sample-level snRNA-seq cell-type proportions and quantitative histological features (fibrous cap thickness, lipid core area, calcification, neovessel area). Heat map colour indicates correlation coefficient; asterisks denote nominal P < 0.05. scRNA-seq, single-cell RNA sequencing; snRNA-seq, single-nucleus RNA sequencing; SMC, smooth muscle cell; EC, endothelial cell; MP, macrophage; EndoMT, endothelial-to-mesenchymal transition; C1Q, complement component 1q; H&E, hematoxylin and eosin; NV, neovessels; CC, calcification; LC, lipid core; FC, fibrous cap; PT, pseudotime; CI, confidence interval; Sym, symptomatic; Asym, asymptomatic; OR, odds ratio; I², I-squared heterogeneity statistic; ρ, Pearson correlation coefficient.

To determine whether symptomatic status was associated with altered cellular state progression, we compared lineage-specific pseudotime distributions using *condiments* (**Supplementary Fig. 3, Supplementary Table 3**), followed by sample-level analyses based on weighted median pseudotime and terminal-state enrichment (**Fig.s 4b, c**). CD4⁺ T cells showed the most consistent evidence of altered state progression in symptomatic plaques. Further, CD4⁺ T cells showed significantly later median pseudotime in snRNA-seq (difference = 0.10, 95% CI = 0.03–0.15) and in the cross-modality meta-analysis (difference = 0.07, 95% CI = 0.02–0.13). Terminal-state occupancy was greater in snRNA-seq (OR = 3.62, 95% CI = 1.68–7.84) and in the meta-analysis (OR = 3.12, 95% CI = 1.65–5.88). Notably, this association was strongest in snRNA-seq despite substantially greater T-cell recovery and finer terminal-state resolution in scRNA-seq, arguing against preferential T-cell recovery as an explanation for the disease association. Osteochondrogenic SMCs showed significantly later weighted median pseudotime in scRNA-seq (difference = 0.13, 95% CI = 0.01–0.21) and in the cross-modality meta-analysis (difference = 0.03, 95% CI = 0.00–0.06), with a concordant shift in snRNA-seq. Terminal-state occupancy was likewise greater in scRNA-seq (OR = 3.47, 95% CI = 1.01–11.98), with a concordant direction of effect in the meta-analysis (OR = 2.53, 95% CI = 0.89–7.21).

EndoMT ECs likewise showed greater terminal-state occupancy in symptomatic plaques in scRNA-seq (OR = 4.82, 95% CI = 1.53–15.13) and in the cross-modality meta-analysis (OR = 3.34, 95% CI = 1.25–8.96). EndoMT ECs were more abundant in snRNA-seq at baseline, whereas the symptom-associated terminal-state shift was strongest in scRNA-seq, again making differential state recovery alone an unlikely explanation. By contrast, CD8⁺ T-cell terminal-state enrichment was restricted to scRNA-seq (OR = 12.37, 95% CI = 1.45–105.5) and was not supported in snRNA-seq (OR = 0.71), resulting in substantial cross-modality heterogeneity (I² = 84%).

We next used matched histopathology from the AtherOMICS cohort to provide an independent phenotypic context for cell-state and transcriptional findings detected by snRNA-seq. We annotated key structural features, including lipid core area, calcification burden, neovessel area, and fibrous cap thickness (**Fig. 4d**). Total, contractile, and osteochondrogenic SMC proportions each correlated with higher fibrous cap thickness (r = 0.63, r = 0.63, and r = 0.56, respectively), whereas the proportion of EndoMT ECs and plasma cells each correlated positively with calcification burden (r = 0.59 and r = 0.58; **Fig. 4e, Supplementary Table 4**). Macrophages showed an interesting pattern, with TREM2⁺ foamy macrophages correlated with a larger lipid core area (r = 0.52) and less calcification burden (r = −0.55), whereas C1Q^+^ remodeling macrophages were associated with a smaller lipid core area (r = −0.54). HMOX1⁺ heme-handling macrophages showed a negative correlation with the proportion of calcification burden (r = −0.54). Finally, the proportion of naïve T cells correlated with a larger lipid core area (r = 0.52). These associations should be interpreted with caution, as although they represent large effects, they did not reach statistical significance after correcting for multiple comparisons, likely due to the small sample size (n = 15-16 per endpoint). Nevertheless, their effect sizes and direction provided orthogonal context for the subsequent vascular wall population findings in the snRNA-seq dataset.

### Symptomatic plaque-associated transcriptional changes are modality-dependent and point to inflammatory and EMT-related programmes

To characterize symptom-associated transcriptional remodeling, we performed cell-type-specific pseudobulk differential expression analyses on scRNA-seq and snRNA-seq datasets. Consistent with the substantial gene-level differences between scRNA-seq and snRNA-seq defined above, symptom-associated differential expression was strongly modality-dependent, with 361 significant DEGs identified in scRNA-seq compared with only 17 in snRNA-seq (**Fig. 5a**). ECs and T cells accounted for the majority of scRNA-seq DEGs (196 and 102 genes, respectively), with *ADAMTS2* (log₂FC = 7.00, FDR = 2.4 × 10⁻³) leading in ECs. Fibroblasts represented the most transcriptionally altered cell population in snRNA-seq (8 genes) (**Fig. 5a**), led by a substantially lower expression of *RGS5* among symptomatic plaques (log₂FC = −4.41, FDR = 1.8 × 10⁻⁴) (**Fig. 5b**), the largest-effect-size gene in the snRNA-seq dataset. *RGS5* expression was also positively associated with fibrous cap thickness, a marker of plaque stability (ρ = 0.716, FDR = 0.016) (**Fig. 5c,d**). Fibroblasts also showed lower *GUCY1A1* (log₂FC = −1.93, FDR = 0.017) and higher *MMP2* (log₂FC = 2.27, FDR = 0.017) and *TIMP3* (log₂FC = 2.68, FDR = 0.007) expression in symptomatic plaques, and SMCs showed higher *COL11A1* (log₂FC = 2.29, FDR = 0.017) in snRNA-seq. Importantly, no genes reached FDR statistical significance in the same cell type across both modalities (**Supplementary Table 5**). Baseline differences in cell-type recovery between modalities did not account for these gene-level associations (**Fig. 2c**).

**Fig. 5.**
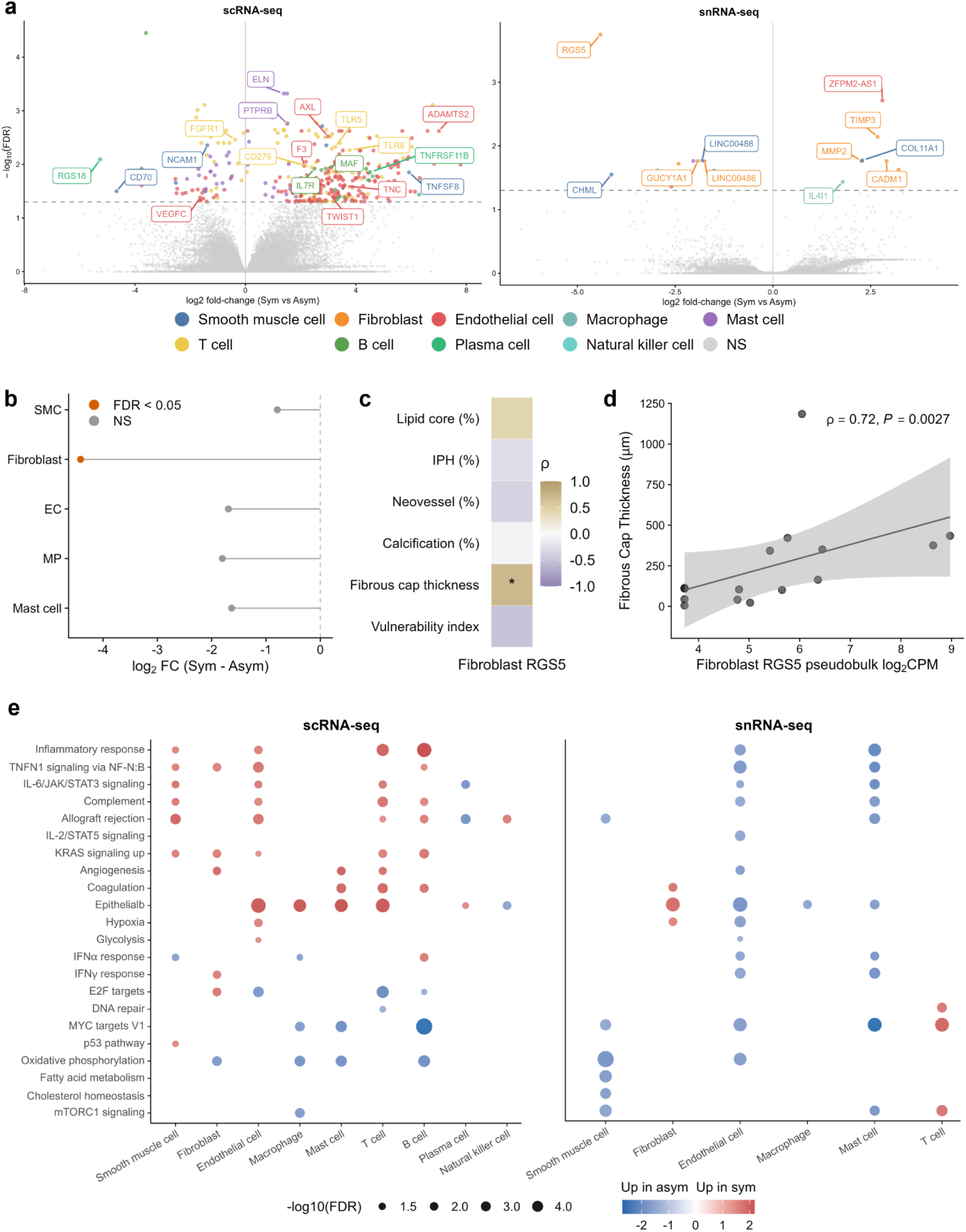
Symptomatic plaque status is associated with modality-dependent gene- and pathway-level remodelling, anchored by fibroblast *RGS5* loss. (a) Volcano plots of cell-type-specific pseudobulk differential expression between symptomatic and asymptomatic plaques in scRNA-seq (left) and snRNA-seq (right). Genes are coloured by the broad cell type in which they reached FDR < 0.05; grey points denote non-significant (NS) genes. Dashed horizontal line marks FDR = 0.05. (b) Cell-type-specific log_2_ fold change (symptomatic − asymptomatic) for *RGS5* in snRNA-seq. Orange, FDR < 0.05; grey, NS. (c) Spearman correlations (ρ) between fibroblast *RGS5* pseudobulk expression and quantitative histological features. Asterisks denote FDR < 0.05. (d) Fibroblast *RGS5* pseudobulk expression (log_2_CPM) versus fibrous cap thickness. Line, linear fit; shading, 95% confidence interval. (e) Bubble plots of Hallmark pathway enrichment in symptomatic versus asymptomatic plaques by broad cell type, in scRNA-seq (left) and snRNA-seq (right). Colour indicates normalized enrichment score; red denotes enrichment in symptomatic plaques and blue denotes enrichment in asymptomatic plaques. Point size indicates −log_10_(FDR). scRNA-seq, single-cell RNA sequencing; snRNA-seq, single-nucleus RNA sequencing; SMC, smooth muscle cell; EC, endothelial cell; MP, macrophage; FDR, false discovery rate; NS, not significant; FC, fold change; Sym, symptomatic; Asym, asymptomatic; IPH, intraplaque hemorrhage; CPM, counts per million; ρ, correlation coefficient; P, P value; IFNα, interferon alpha; IFNγ, interferon gamma; TNFα, tumor necrosis factor alpha; NF-κB, nuclear factor kappa B; IL-2, interleukin 2; STAT5, signal transducer and activator of transcription 5; mTORC1, mechanistic target of rapamycin complex 1; UV, ultraviolet

Since gene-level differential expression showed limited concordance between modalities, we next examined Hallmark pathway-level changes (**Fig. 5e; Supplementary Table 6**). In scRNA-seq, symptomatic plaques showed enrichment of EMT-related and inflammatory gene sets: EMT in ECs (NES = 2.03, FDR = 5.0 × 10⁻¹¹) and T cells (NES = 2.05, FDR = 2.0 × 10⁻⁹), TNFα–NFκB signaling in ECs (NES = 1.73, FDR = 7.1 × 10⁻⁵; alongside IL6–JAK– STAT3 and complement), and inflammatory response in T cells (NES = 1.93, FDR = 9.1 × 10⁻⁷). In snRNA-seq, fibroblasts showed EMT enrichment in symptomatic plaques (NES = 1.96, FDR = 2.2 × 10⁻⁵), alongside RGS5 downregulation. By contrast, most snRNA-seq pathways were enriched in asymptomatic plaques, most prominently in ECs, including EMT (NES = −1.99, FDR = 6.9 × 10⁻⁶), inflammatory response, hypoxia and glycolysis. Since these EC pathways overlap with stress-associated responses, and ECs showed a significant modality-by-symptom interaction for a dissociation-stress gene set (FDR = 3.0 × 10⁻⁵; **Supplementary Table 7**), EC pathway directionality should be interpreted cautiously.

### Genetic risk for atherosclerotic disease could be driven by genes differentially expressed in symptomatic plaques

Finally, we integrated the scRNA-seq and snRNA-seq datasets with genome-wide association study (GWAS) data for coronary artery disease (CAD), large-artery atherosclerotic stroke (LAS), peripheral artery disease (PAD), and carotid plaque presence (CaP) toTo identify plaque cell types and states implicated by genetic risk and to determine whether genetically prioritized genes were differentially expressed between symptomatic and asymptomatic plaques, we integrated the scRNA-seq and snRNA-seq datasets with GWAS data for coronary artery disease (CAD), large artery stroke (LAS), peripheral artery disease (PAD), and carotid plaque presence (CaP).

We first used stratified linkage disequilibrium score regression (S-LDSC) to test whether disease heritability was enriched in genomic annotations derived from specific plaque cell types and states.^19^ S-LDSC revealed strong enrichment of genetic risk in vascular wall cell types. Fibromyocyte, contractile, and osteochondrogenic SMC states, together with fibroblasts, were significantly enriched for CAD and PAD heritability in both scRNA-seq and snRNA-seq datasets (FDR < 0.05; **Fig. S4a**), with CaP showing a similar pattern in snRNA-seq. Furthermore, there was enrichment of quiescent ECs for CAD in both modalities, whereas proangiogenic and EndoMT EC states were enriched in scRNA-seq for CAD, PAD, and LAS. Alternative approaches prioritizing risk genes from GWAS signals based on either colocalization with expression quantitative trait loci (eQTLs) in arterial tissue or the inclusive locus-to-gene (L2G) model by Open Targets confirmed similar enrichment in vascular wall cells, mainly SMC states (**Supplementary Fig. 4b**, **Supplementary Tables 8-10)**.

Next, we investigated whether individual genes implicated in the genetic risk of atherosclerotic disease were differentially expressed between symptomatic and asymptomatic plaques (**Fig. 6a and Supplementary Fig. 4c**; **Supplementary Table 11**). Focusing on genes supported by colocalization between arterial tissue expression and atherosclerotic disease risk, we identified 12 genes that were differentially expressed in specific cell types between symptomatic and asymptomatic individuals. Among them, *TWIST1* (log₂FC = 3.04), *TMEM59L* (log₂FC = 4.25), *BMP1* (log₂FC = 1.10), and *PCDHGB5* (log₂FC = 1.19) were upregulated in ECs of symptomatic plaques, whereas *ATP2B1-AS1* was downregulated (log₂FC = −1.05) in ECs of symptomatic plaques. *TMEM59L*, *MRAS* and *HLA-DQB2* were additionally upregulated in T cells, while *GUCY1A1* was downregulated in MPs in scRNA-seq and in fibroblasts in snRNA-seq. Notably, *TWIST1* showed strong colocalization of aortic expression with all four atherosclerotic phenotypes (CAD posterior probability hypothesis 4 [PP.H4] = 0.994, LAS = 0.994, PAD = 0.985, CaP = 0.873; **Fig. 6b**), whereas, *MRAS* showed colocalization of arterial expression with CAD risk across all three tested tissues (aorta PP.H4 = 0.996, coronary PP.H4 = 0.997, tibial PP.H4 = 0.996), but not with LAS, PAD, or CaP (**Fig. 6c**). Using an alternative prioritization strategy based on L2G scores, we observed partially overlapping associations, including *BMP1* and *GUCY1A1*, while identifying additional DEGs mainly in ECs, including *CELSR2* (log₂FC = 6.60), *NGF* (log₂FC = 5.54), *EDNRA* (log₂FC = 4.81), *ITIH4* (log₂FC = 4.04), *SFRP2* (log₂FC = 3.59), and *FST* (log₂FC = 3.47) (**Fig. S4c**).

**Fig. 6.**
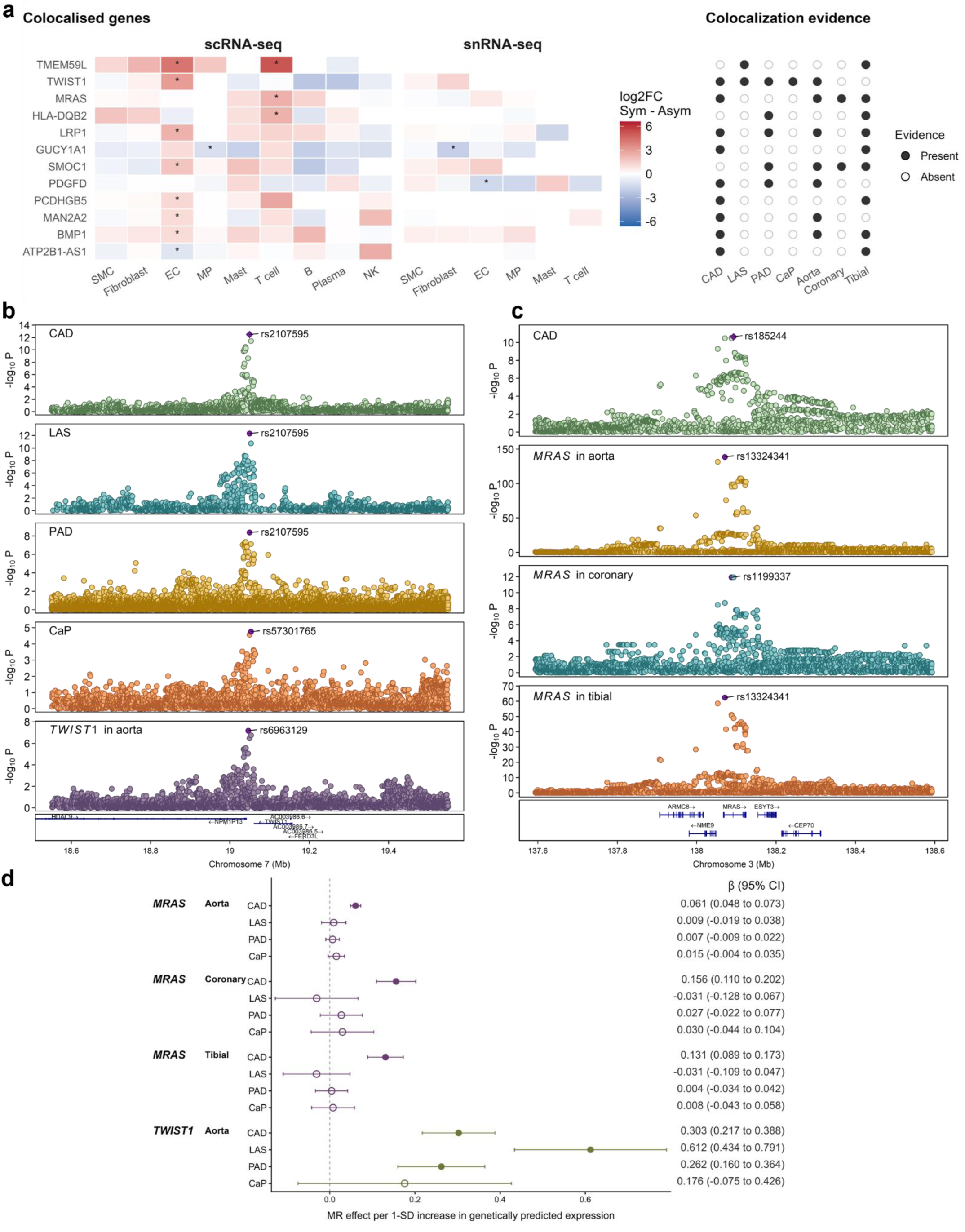
Colocalized genes converge on symptomatic-plaque differential expression at endothelial *TWIST1* and T-cell *MRAS*. (a) Left, cell-type-specific log_2_ fold change (symptomatic versus asymptomatic) for genes identified through colocalization (coloc.abf, PP.H4 > 0.8) between CAD, LAS, PAD, or CaP GWAS and GTEx aorta, coronary, or tibial artery eQTL, in scRNA-seq and snRNA-seq. Asterisks mark cell types in which the gene reached FDR < 0.05. Right, evidence matrix indicating colocalization support for each gene across CAD, LAS, PAD, CaP and each GTEx artery. (b) Colocalization of *TWIST1* expression in GTEx aorta with CAD, LAS, PAD and CaP GWAS signal at 7p21.1. Purple diamond, index SNP. (c) Colocalization of MRAS expression in GTEx aorta, coronary, and tibial arteries with CAD GWAS signal at 3q22.3. Purple diamond, index SNP. (d) Cis-Mendelian randomization effect estimates (β, 95% CI) for genetically predicted *MRAS* and *TWIST1* expression on CAD, LAS, PAD and CaP risk. Filled circles, q < 0.05; open circles, non-significant. scRNA-seq, single-cell RNA sequencing; snRNA-seq, single-nucleus RNA sequencing; SMC, smooth muscle cell; EC, endothelial cell; MP, macrophage; NK, natural killer; FC, fold change; Sym, symptomatic; Asym, asymptomatic; CAD, coronary artery disease; LAS, large artery stroke; PAD, peripheral artery disease; CaP, carotid plaque; MR, Mendelian randomization; SD, standard deviation; CI, confidence interval; β, beta coefficient; rs, reference SNP identifier

To further evaluate the genetic support and directionality of these associations, we performed cis-Mendelian randomization (MR) analyses (**Supplementary Table 12**). These analyses showed concordant effects for genetically proxied expression of several genes. Higher genetically proxied *TWIST1* expression in aorta was associated with higher LAS risk (β = 0.612, q = 1.4 × 10⁻⁹), CAD risk (β = 0.303, q = 2.5 × 10⁻¹¹), and PAD risk (β = 0.262, q = 1.8 × 10⁻⁵), while higher genetically proxied *MRAS* expression in aorta, tibial artery, and coronary artery was associated with increased CAD risk (β = 0.061, q = 1.5 × 10⁻²¹ in aorta; β = 0.131, q = 9.5 × 10⁻⁹ in tibial artery; and β = 0.156, q = 2.7 × 10⁻¹⁰ in coronary artery) (**Fig. 6d**). Higher genetically proxied *HLA-DQB2* expression in tibial artery was associated with increased PAD risk (β = 0.047, q = 1.3 × 10⁻⁷), whereas lower genetically proxied *GUCY1A1* expression in tibial artery was associated with increased CAD risk (β = −0.419, q = 6.6 × 10⁻¹³).

## DISCUSSION

In this study, we compared cellular and molecular features between symptomatic and asymptomatic human atherosclerotic plaques using snRNA-seq and scRNA-seq data across 53 patients. We found that symptomatic plaques exhibit a depletion of fibroblasts characterized by reduced *RGS5* expression, cellular shifts toward osteochondrogenic SMC, CD4⁺ T-cell, and EndoMT states, and enrichment of inflammatory and EMT-related transcriptional programs in ECs, T cells, and fibroblasts. Integration of cell-specific transcriptional signatures of symptomatic plaques with genomic risk data for atherosclerotic disease nominated local *TWIST1*, *GUCY1A1*, and *MRAS* expression as candidate drivers of symptomatic plaque transformation.

Fibroblasts showed both compositional and transcriptional remodeling in symptomatic plaques. SnRNA-seq profiling revealed significant fibroblast depletion, an association that remained significant in the cross-modality meta-analysis. Previous single-cell and experimental studies have identified distinct plaque fibroblast states and shown that fibroblast-like cells can accumulate within the fibrous cap and influence cap thickness.^20–22^ In particular, IL-1β inhibition in models of clonal hematopoiesis increased fibroblast-like cell accumulation and fibrous cap thickness, whereas depletion of these cells reduced cap thickness.^22^ The remaining fibroblasts in symptomatic plaques showed pronounced transcriptional remodeling, including lower *RGS5* and *GUCY1A1* and higher *MMP2* and *TIMP3* expression. These changes were preferentially detected in snRNA-seq. Notably, experimental studies have also shown that RGS5 loss perturbs the vascular niche through complementary mechanisms: reduced *RGS5* promotes endothelial NF-κB signalling and dysfunction in human endothelial cells,^23^ whereas pericyte-specific *Rgs5* loss in mice induces TGFβ2-dependent paracrine fibroblast activation.^24^ A broader role in plaque remodeling is supported by apolipoprotein E-deficient mice, in which *Rgs5* deficiency increased lesion and necrotic-core burden while reducing SMC and collagen content.^25^ In our cohort, higher fibroblast *RGS5* expression was associated with greater fibrous cap thickness. Together, these findings place the observed *RGS5* downregulation in the context of experimental studies linking RGS5 deficiency to NF-κB–mediated inflammatory signalling, fibroblast activation and reduced plaque stability. Fibroblasts in symptomatic plaques were further characterized by reduced *GUCY1A1* expression, which converged with genomic data, pointing to arterial downregulation of *GUCY1A1* expression as a driver of CAD risk. *GUCY1A1* encodes the alpha-1 subunit of soluble guanylate cyclase (sGC), which is the primary receptor driving the vasoprotective actions associated with nitric oxide signaling.^26^ Experimental studies indicate that the consequences of altered sGC signalling in atherosclerosis are strongly cell-context dependent. Functional characterization of the *GUCY1A3/GUCY1A1* CAD locus implicated altered sGC signalling in vascular SMC migration,^27^ whereas platelet-specific loss of sGC increased plaque formation and vascular inflammation and pharmacological sGC stimulation reduced leukocyte recruitment and atherosclerosis.^28^ These divergent effects argue against a uniform vascular consequence of sGC perturbation. The concomitant increase in *MMP2* and *TIMP3* further suggests active extracellular-matrix turnover; experimental studies showing reduced lesion development with MMP2 deficiency and aggravated atherosclerosis with TIMP3 loss support functional relevance of this protease–inhibitor axis, although increased TIMP3 in symptomatic fibroblasts may represent a compensatory response.^29,30^

We also found a shift toward an osteochondrogenic SMC state in symptomatic plaques. In scRNA-seq, osteochondrogenic SMCs from symptomatic plaques were further along the pseudotime trajectory and showed greater representation of terminal states; snRNA-seq showed the same directional shift. Consistent with this finding, COL11A1, a cartilage-associated collagen, was increased in the broad SMC compartment. Lineage-tracing and single-cell studies have linked fibromyocyte states to fibrous cap formation and lower CAD risk, whereas osteochondrogenic states are consistently associated with cartilage-like matrix remodeling and calcification.^9,10^ Consistent with our findings, osteogenic and extracellular-matrix programs have been reported in advanced symptomatic human plaques,^31^ and CAD-associated regulatory activity has been localized to SMC transition states.^32^

Another significant difference was the higher representation of terminal EndoMT-associated EC states in symptomatic plaques. ECs also showed enrichment of inflammatory and EMT-associated pathways, including TNF–NFκB, IL6–JAK–STAT3, complement activation, hypoxia, and glycolysis. EndoMT has been described as a feature of advanced and unstable atherosclerotic plaques,^33^ but its effects appear to depend on the specific molecular program involved.^34^ Recent experimental work has identified TWIST1 as a driver of endothelial phenotypic modulation and EndoMT in atherosclerosis, with endothelial *Twist1* deletion reducing lesion size and promoting a more stable plaque phenotype.^35^ Notably, *TWIST1* was also upregulated in ECs from symptomatic plaques in our study. This finding was further supported by genetic analyses, which linked variants associated with increased *TWIST1* expression in human arterial tissue to atherosclerotic phenotypes across vascular beds, including CAD, LAS, PAD, and CaP. Together, these findings identify endothelial *TWIST1* as a potential mediator of EMT-associated changes in symptomatic plaques.

In line with the phenotypic differences between symptomatic and asymptomatic plaques, which were most prominent in vascular wall cells, genetic analyses also highlighted these cell types. S-LDSC, L2G, and arterial eQTL-based analyses, genetic risk for CAD, LAS, PAD, and CaP was particularly enriched in contractile, fibromyocyte, and osteochondrogenic SMC states and in fibroblasts, with additional endothelial associations depending on the phenotype and modality. These findings were consistent across complementary approaches to cell-state enrichment and gene prioritization and were observed across both scRNA-seq and snRNA-seq datasets. They are also consistent with human single-nucleus chromatin studies that have localized CAD-associated regulatory activity to phenotypically modulated SMC states, including fibromyocytes.^32^

The most reproducible immune-state difference involved CD4⁺ T cells. In snRNA-seq, CD4⁺ cells occupied later pseudotime positions and showed greater terminal-state commitment than in asymptomatic plaques. This is consistent with previous human carotid studies reporting greater CD4⁺ T-cell activation and differentiation in symptomatic lesions and plaque-specific clonal expansion with evidence of recent T-cell receptor engagement among effector CD4⁺ cells,^8,15^ supporting local antigen-experienced differentiation. *MRAS*, which was upregulated in symptomatic T cells here and strongly supported by CAD colocalization and cis-MR, provides a potential genetic link to this phenotype. MRAS forms a SHOC2–PP1C complex that promotes RAF–MAPK signaling,^36^ although CAD-associated regulatory effects at this locus have previously been localized predominantly to SMC transition states, including fibromyocytes.^32^ Together, these findings extend earlier descriptions of plaque T-cell activation by identifying a sample-level CD4⁺ differentiation shift associated with symptomatic disease and nominate *MRAS* as a potential link between genetic risk and this immune-state transition.

Previous studies of human atherosclerosis have used single-cell technologies to define plaque-cell heterogeneity and disease-associated states,^37^ but direct comparisons of how whole-cell and nuclear RNA profiling capture these states have been limited.^38,39^ In this study, mean normalized expression showed only modest concordance between scRNA-seq and snRNA-seq. These differences likely reflect differences in the cellular compartments captured by the two assays, as well as differences in cell recovery and transcriptional responses to tissue dissociation. Comparisons in other tissues have shown similar differences between scRNA-seq and snRNA-seq in pre-mRNA representation, cell-type recovery, and responses to tissue processing.^38,39^ Modality-specific findings remain informative when the corresponding modality has a clear measurement advantage for the relevant biological compartment.

Concordance between the two modalities was greater for broad cell identities than for individual genes or pathways. At the disease-association level, no cell-specific DEGs reached significance in both modalities, and EC pathway effects differed in direction. Cross-modality meta-analysis supported fibroblast depletion and shifts toward terminal CD4⁺ T-cell and later osteochondrogenic SMC states in symptomatic plaques, although individually each of these effects reached significance in only one modality. The cross-modality analyses therefore serve as an interpretive framework for the disease associations rather than a separate benchmarking endpoint, and modality-specific associations should be interpreted cautiously.

Several limitations should be considered. First, although samples from 53 patients were included in the overall analyses, many comparisons remained underpowered, limiting our ability to detect anything but relatively large cellular and molecular differences. This was particularly relevant for cell-abundance comparisons, where fibroblast depletion was the only significant difference identified, and for correlations with histology, which were restricted to in-house snRNA-seq data from only 15-16 patients per endpoint. Larger datasets, ideally integrated across centers, will be important to enable better-powered comparisons. Second, we observed substantial differences in both cellular composition and transcriptional profiles between scRNA-seq and snRNA-seq, reflecting inherent differences in the strengths and limitations of the two technologies, including differential recovery of cytoplasmic and nuclear transcripts, dissociation-induced stress signatures in scRNA-seq, and cell-type recovery biases. These differences introduce technical variability that may have obscured biological differences between symptomatic and asymptomatic plaques. Accordingly, differences in the number of differentially expressed genes between modalities should not be interpreted as differences in sensitivity. Third, the scRNA-seq cohort comprised four independently generated public datasets, whereas the snRNA-seq cohort was generated independently within AtherOMICS, and the two modalities were obtained from different samples. Thus, modality effects cannot be fully separated from cohort- and study-specific effects, limiting direct comparisons between the two datasets. Fourth, the symptomatic–asymptomatic comparisons were necessarily cross-sectional and therefore cannot establish whether the identified cellular states precede clinical events, contribute to plaque destabilization, arise as consequences of advanced disease, or represent compensatory responses. Similarly, pseudotime analyses infer transcriptional ordering from static measurements and cannot establish temporal progression or lineage fate in vivo.^40^

In conclusion, this work provides a comprehensive atlas of cellular and molecular differences between symptomatic and asymptomatic human atherosclerosis. Our findings highlight fibroblast depletion, cellular shifts toward osteochondrogenic SMC, EndoMT, and CD4⁺ T-cell states, and molecular changes across cell types pointing to inflammatory and EMT-associated transcriptional programmes as signatures of symptomatic plaques. Integration with genetic risk data nominated locally DEGs, including *TWIST1*, *GUCY1A1*, and *MRAS*, as candidate causal drivers of symptomatic plaque transformation. These findings identify specific cellular programs that distinguish symptomatic from asymptomatic human carotid plaques and provide cell-specific prioritized targets for mechanistic studies of plaque destabilization.

## METHODS

### Study cohort for single-nucleus RNA-seq data

AtherOMICS is a prospective biobank enrolling patients undergoing carotid or femoral endarterectomy at the Vascular Surgery and Neurology Departments, LMU University Hospital Munich. Eligible participants were consecutively identified by screening of admission and surgery records; inclusion required age ≥18 years, endarterectomy scheduled within 7 days, and provision of written informed consent. Exclusion criteria were inability to consent, restenosis after prior endarterectomy or stenting, and stenosis attributable to prior local radiation. The study was approved by the Ethics Committee of the Faculty of Medicine, Ludwig-Maximilians-Universität Munich (protocol numbers 121-09, 22-0135, 23-0772), conformed to the 2008 Declaration of Helsinki, and all participants provided written informed consent.

Carotid stenosis was classified as symptomatic or asymptomatic based on a qualifying ipsilateral cerebrovascular event including ischemic stroke, transient ischemic attack, amaurosis fugax, or central retinal artery occlusion. Plaque tissue was collected intraoperatively and dissected into 5 mm segments centered on the region of greatest atherosclerotic burden, defined by plaque thickness; surrounding segments were flash-frozen and stored at −80 °C for omics profiling. Segment location (proximal versus distal) was retained as a covariate in downstream analyses.

### Single-cell and single-nucleus RNA-seq data processing and integration

Publicly available scRNA-seq datasets of human carotid atherosclerotic plaques were identified by searches of PubMed and the Gene Expression Omnibus from database inception to 9 June 2025. Searches combined terms relating to human carotid atherosclerotic plaques and carotid endarterectomy, scRNA-seq, and symptomatic status. Candidate records were screened for the following exclusion criteria: datasets not comprising human carotid plaque tissue, datasets without individual-level symptomatic or asymptomatic clinical annotation, and datasets without reprocessable carotid-plaque expression data. Of 16 records identified, 11 represented unique studies or datasets and four met the inclusion criteria, comprising 36 samples (16 symptomatic and 20 asymptomatic; **Supplementary Table 13**).^8,10,20,31^ Mocci et al. provided Smart-seq2 gene-by-cell read-count matrices, whereas the remaining datasets provided UMI-based count matrices. All datasets were processed using a uniform Seurat-based workflow (v5.0.3).^41^ Cells were retained if they contained >300 detected genes, >1,000 counts, and <20% mitochondrial reads. Each dataset was normalized using SCTransform with mitochondrial read fraction regressed out, followed by principal component analysis (PCA). Doublets were identified within each dataset using *DoubletFinder*,^42^ with an expected doublet rate of 5%, and only singlets were retained. Highly variable genes (3,000 per dataset) were selected using the vst method, and integration features were selected across datasets using SelectIntegrationFeatures. Dataset integration and batch correction were performed using *Harmony* (v1.2.3)^43^ on the PCA embedding, accounting for sample identity and sequencing technology. The Harmony-corrected embedding (dimensions 1–10) was used to construct a shared nearest-neighbor graph (k = 10), and clustering was performed using the Louvain algorithm at a resolution of 1.3.

Sample collection, nuclei isolation, library preparation, sequencing, read alignment, and UMI quantification for human carotid plaque snRNA-seq have been described previously.^12^ For the present study, raw UMI count matrices and corresponding gene-barcode annotation files were imported into R and processed using *Seurat*. Nuclei were retained if they had >150 detected genes, >150 total UMIs, and <20% of UMIs mapping to mitochondrial genes. After quality control, doublets were identified and removed on a per-sample basis using *DoubletFinder* (v2.0.4), assuming an expected doublet rate of approximately 4% per library. Gene expression was normalized and variance-stabilized using SCTransform, with the mitochondrial fraction regressed out to reduce technical variation. Highly variable features (1,500 genes) were selected across samples, integration anchors were identified using FindIntegrationAnchors, and a shared integrated assay was generated with IntegrateData. PCA was performed on the integrated assay, and the first 15 principal components were used to construct a shared nearest-neighbor graph (k = 40). Unsupervised clustering was performed using the Louvain algorithm with a resolution of 1.4.

### Cell type annotation

Broad lineage identities were assigned using canonical marker genes obtained from published single-cell studies and atlases of human atherosclerotic and vascular tissues.^9,11,13–18^ Cluster-level expression profiles from SCTransform-normalized data were evaluated using three complementary approaches: marker-based scoring, reference-based correlation using *clustifyr* (v1.18.0), and gene set enrichment analysis using *GSVA/ssGSEA* (v2.0.7). Final lineage labels were determined based on concordance across these approaches, together with manual inspection of established marker gene expression. Finer transcriptional subtypes were resolved using a hierarchical annotation strategy. Clusters assigned to each broad lineage were subset and re-analyzed independently to identify within-lineage states, which were subsequently propagated back to the full dataset.

### Cross-modality cell-type discriminability analysis

To assess whether broad cell-type annotations were transcriptionally comparable across scRNA-seq and snRNA-seq, we quantified within- and cross-modality cell-type discriminability using MetaNeighbor.^44^ Expression matrices were restricted to genes shared by at least 10 cells or nuclei in either modality. To limit differences in cell-type abundance, each broad cell type was downsampled to a maximum of 2,000 cells or nuclei per modality. Counts were library-size normalized and log-transformed, and highly variable genes were identified using MetaNeighbor::variableGenes with modality specified as the study variable. Cell-type discriminability was quantified using MetaNeighborUS, with modality as the study identifier and harmonized broad cell type as the cell-type label. Hierarchical clustering of AUROC profiles was performed using Euclidean distances and complete linkage.

### Compositional analysis

Cell-type proportions were compared between scRNA-seq and snRNA-seq using *propeller* (*speckle* v1.6.0, logit transformation).^45^ For each cell type, sample-level proportions were logit-transformed and modeled by modality, yielding empirical Bayes-moderated t-statistics and modality-specific mean proportions. Analyses were performed at both broad and fine annotation levels; P values were adjusted separately within each level using the Benjamini-Hochberg (BH) method.

Associations between cell-type composition and symptomatic status were evaluated separately for each modality using beta-binomial regression with a logit link, with cell-type counts modeled relative to total cells recovered per sample (asymptomatic as reference). In the snRNA-seq dataset, generalized linear models included clinical group, plaque segment location, centered and scaled age, and sex as fixed effects, implemented in *glmmTMB* (v1.1.11). In the integrated scRNA-seq dataset, generalized linear mixed models included clinical group and sex as fixed effects and a random intercept for dataset of origin. Finer subtype models used the corresponding broad parent population as the denominator rather than total cells per sample. P values were adjusted within each modality using the BH method.

To derive cross-modality estimates, logit-scale effect sizes and standard errors from scRNA-seq and snRNA-seq were combined using random-effects meta-analysis using *metafor* (v4.8.0), restricted to cell types tested in both modalities. Heterogeneity was quantified using I². Meta-analytic P values were adjusted using the BH method, applied separately at each annotation level.

### Pseudotime and lineage trajectory analysis

Pseudotime and lineage reconstruction were performed separately for each broad lineage to characterize transcriptional trajectories among identified subtypes. UMAP embeddings were used as input to *Slingshot* (v2.14.0)^46^ for trajectory inference in both modalities, with Seurat cluster assignments specified as lineage nodes, and biologically informed root clusters were selected for each lineage.

Differences between asymptomatic and symptomatic plaques along inferred trajectories were evaluated using the *condiments* package (v1.14.0).^47^ Lineage-level differences in pseudotime distributions were tested using progressionTest, and lineage endpoint usage differences were assessed using fateSelectionTest, with both global and pairwise comparisons. P values were adjusted for multiple testing within each lineage run using the BH method.

Donor-level pseudotime progression was computed as the weight-averaged median normalized pseudotime per sample per branch, using *Slingshot* curve weights as the weighting factor. Differences between symptomatic and asymptomatic samples were quantified using the Hodges-Lehmann estimator of the median shift in weighted-median pseudotime, computed separately for each lineage branch; standard errors were derived from 2,000 bootstrap resamples drawn independently within each clinical group, with branches tested only when at least three samples per group were available. Cross-modality effect sizes were obtained using fixed-effects meta-analysis of scRNA-seq and snRNA-seq Hodges-Lehmann estimates using *metafor* (v4.8.0), restricted to branches tested in both modalities; heterogeneity was quantified using I².

Terminal state occupancy was defined for each lineage branch after pseudotime was normalized to the [0,1] interval. Cells or nuclei were assigned to the branch with the highest *Slingshot* curve weight, and terminal occupancy was defined as the fraction of assigned cells or nuclei with normalized pseudotime >0.7, computed at the sample-branch level for samples that contributed at least 10 cells or nuclei to that branch. Branches were tested when they contained at least three samples per clinical group. Differences between symptomatic and asymptomatic plaques were evaluated using beta-binomial models with the number of terminal and non-terminal cells or nuclei as the response variable; snRNA-seq models included clinical group and sex as fixed effects, and scRNA-seq models included clinical group and sex as fixed effects, with dataset of origin as a random intercept. Cross-modality effect sizes were obtained via a fixed-effects meta-analysis of logit-scale estimates from both modalities using *metafor*, restricted to branches tested in both modalities. P values were adjusted using the BH method within each modality.

### Cross-modality gene expression concordance analysis

To compare gene expression between scRNA-seq and snRNA-seq at both the global and cell-type-specific levels, pseudobulk count matrices were generated within each modality by aggregating raw counts across cells or nuclei within each sample (global) or within each sample and annotated broad cell type (cell-type-specific). Analyses were restricted to genes shared between modalities. Lowly expressed genes were filtered by retaining genes with mean counts per million (CPM) ≥ 0.5 and detectable expression (CPM > 0) in at least 10% of pseudobulk profiles; within each analysis (global or cell-type-specific), genes were then further filtered using edgeR::filterByExpr given the corresponding design matrix.^48^ Library sizes were subsequently normalized using Trimmed Mean of M-values (TMM) normalization, and normalized expression values were obtained in voom space. Mean logCPM was calculated separately for scRNA-seq and snRNA-seq by averaging across pseudobulk profiles within each modality. Gene-wise mean expression levels in scRNA-seq and snRNA-seq were then compared using global and cell-type-specific scatter plots, and concordance between modalities was summarized using the squared Pearson correlation coefficient (R²).

### Differential expression analysis

Cell-type-specific differential expression was assessed using a pseudobulk framework. Raw counts were aggregated per sample and cell type to generate sample-level expression profiles. Genes with insufficient expression were excluded by retaining genes with mean CPM ≥ 0.5 and detectable expression (CPM > 0) in at least 10% of pseudobulk profiles, and additional expression filtering was performed within each cell type using edgeR::filterByExpr given the corresponding design matrix. Pseudobulk profiles with low library sizes were removed within each cell type before testing.

In the snRNA-seq dataset, cell-type-specific differential expression between symptomatic and asymptomatic plaques was assessed using edgeR negative binomial generalized linear model including clinical group as the primary predictor and plaque segment location, centered age, and sex as covariates. Analyses were performed only for cell types with at least three pseudobulk profiles per clinical group and with ≥50 genes remaining after expression filtering. Quasi-likelihood F-tests were used as the primary inference procedure; for cell types in which no genes reached FDR < 0.05, the analysis automatically reverted to likelihood-ratio testing. Multiple testing correction was performed using the BH method, with FDR < 0.05 considered significant.

In the integrated scRNA-seq dataset, differential expression was assessed using edgeR quasi-likelihood F-tests only. For each cell type, design matrices were constructed adaptively to include clinical group and to include sex and/or dataset-of-origin covariates only when these varied within that cell type; models were fitted only when both clinical groups were represented by at least two pseudobulk profiles. P values were adjusted using the BH method, with FDR < 0.05 considered significant.

### Pathway enrichment analysis

To quantify systematic modality-associated shifts in pathway activity between scRNA-seq and snRNA-seq, Hallmark gene sets (MSigDB collection H) were obtained using the *msigdbr* package (v25.1.1).^49^ For each broad cell type, gene-level modality effects were estimated from joint voom models that included modality, clinical group, sex, and their interaction terms, and extracted as the log fold change for scRNA-seq relative to snRNA-seq. These gene-level effects were aggregated within each Hallmark pathway by computing the mean log-fold change across all retained genes after expression filtering and cross-modality gene intersection, restricting analyses to pathways with at least 20 genes represented.

To assess symptom-associated pathway enrichment within each modality, gene set enrichment analysis was performed separately for each broad cell type using Hallmark gene sets. For each cell type, genes from the symptomatic versus asymptomatic pseudobulk differential expression analysis were ranked by log fold change, and enrichment was tested using the *fgsea* (v1.32.4)^50^ multilevel algorithm. Gene set sizes were restricted to 3–1000 genes. Normalized enrichment scores and adjusted P values were reported, with statistical significance defined at FDR < 0.05 for both scRNA-seq and snRNA-seq.

### Subcellular localization enrichment analysis

To determine whether modality-associated transcriptional differences reflected differential capture of nuclear versus cytoplasmic transcript pools, genes were classified using Gene Ontology cellular-component (GO:CC) annotations retrieved via *org.Hs.eg.db*. Nuclear identity was defined by annotation to spliceosome-, transcription factor complex-, or nucleolus/nuclear-body-related terms (eight GO:CC terms); cytosolic identity was defined by annotation to ribosomal and cytosolic ribonucleoprotein complex terms (five GO:CC terms). Genes annotated to both categories were excluded, and analyses were restricted to genes with an unambiguous nuclear or cytosolic annotation.

Modality-associated differential expression was derived from the joint voom model described above (modality × clinical group + sex), using the modality coefficient (scRNA-seq relative to snRNA-seq) from each cell-type-specific fit. Genes with P values < 0.05 and positive log fold change were classified as scRNA-seq-upregulated; genes with P values < 0.05 and negative log-fold change were classified as snRNA-seq-upregulated. For each broad cell type, enrichment of scRNA-seq-upregulated genes among cytosolic annotations and of snRNA-seq-upregulated genes among nuclear annotations was tested using two-sided Fisher’s exact tests. ORs and 95% CIs were calculated with a Haldane-Anscombe correction (addition of 0.5 to all cell counts) when any contingency table cell was zero. P values were adjusted across cell types and tests using the BH method.

### Dissociation stress analysis

To evaluate dissociation stress-related transcriptional differences between scRNA-seq and snRNA-seq, pseudobulk expression profiles were generated for each modality using final broad cell-type annotations. Raw counts were aggregated per sample and cell type, restricted to genes shared between modalities, and filtered to retain genes meeting expression-based criteria and passing filterByExpr with the corresponding design matrix. Pseudobulk counts were normalized using TMM values, transformed using the voom procedure, and analyzed using linear models implemented in the *limma* framework (v3.62.2).^51^ For each broad cell type, scRNA-seq and snRNA-seq pseudobulk profiles were jointly modeled using a design that included modality, clinical group (symptomatic versus asymptomatic), sex, and the modality-by-symptom interaction term, followed by empirical Bayes moderation.

A predefined dissociation/stress gene set comprising immediate early and stress-response genes was evaluated in two complementary ways. First, modality-associated shifts in the stress program were summarized descriptively within each cell type by extracting the modality coefficient (scRNA-seq relative to snRNA-seq) from the joint model and computing the mean log-fold change (scRNA-seq − snRNA-seq) across genes in the stress gene set. Standard errors and 95% confidence intervals were derived from the dispersion of gene-level modality coefficients within the gene set. Second, to formally test whether stress-related differences between symptomatic and asymptomatic plaques were modality-dependent, the modality-by-symptom interaction term was assessed using *limma*’s competitive gene-set test (camera) applied to the interaction contrast.^52^ This test was performed within each broad cell type when sufficient stress-module genes were represented. P values from the interaction gene-set tests were adjusted across cell types using the BH procedure, with FDR-adjusted values reported.

### Association analyses of snRNA-seq cell-state features with quantitative plaque morphology

Plaque histological measures used in histology-integrated analyses were derived from the systematic histological characterization described previously.¹² Briefly, the most diseased plaque segment was fixed, decalcified, paraffin-embedded, and serially sectioned for H&E and Picrosirius red staining. Whole-slide images were analyzed in QuPath,^53^ with plaque features including lipid core, calcification, neovascularization and intraplaque hemorrhage annotated by experienced investigators blinded to sample identity. Primary endpoints comprised lipid-core area, calcification burden, and neovessel area, expressed as a percentage of plaque area, and fibrous cap thickness (µm). For analyses of fibroblast *RGS5* expression, intraplaque hemorrhage (IPH; percentage of plaque area) and plaque vulnerability index were additionally evaluated. The vulnerability index (range, 0–4) was defined as the sum of four binary features: thin fibrous cap (≤25th percentile), large lipid core (≥75th percentile), high IPH burden (≥75th percentile) and the presence of a surface event (fibrous cap rupture or erosion). We derived percentile thresholds from samples with available data for each feature. Since the index incorporates histological features analyzed individually, associations across endpoints are not independent. Analyses included up to 16 samples, with endpoint-specific complete-case sample sizes of 15–16 (n = 15 for fibrous cap thickness and the vulnerability index).

Donor-level proportions of broad and fine cell populations were calculated relative to total nuclei and correlated with each of the four primary histological endpoints using Pearson correlation. P values were adjusted using the Benjamini-Hochberg method. Fibroblast *RGS5*, the largest-effect DEG identified in snRNA-seq, was correlated with each of the six histological endpoints using Spearman rank correlation. Expression was quantified as sample-level fibroblast pseudobulk log₂CPM [TMM-normalized; restricted to samples with ≥10 fibroblast nuclei]. P values were adjusted across the six endpoints using the Benjamini-Hochberg method.

### GWAS and eQTL data sources and preparation

GWAS summary statistics for CAD were obtained from the CARDIoGRAMplusC4D 1000 Genomes-based meta-analysis (113,937 cases, 339,115 controls).^54^ Summary statistics LAS were obtained from the GIGASTROKE trans-ancestry meta-analysis (9,219 LAS cases among 1,503,898 controls).^55^ Summary statistics for PAD were obtained from the Million Veteran Program GWAS (31,307 cases, 211,753 controls, European, African, and Hispanic ancestry).^56^ Summary statistics for CaP were obtained from a GWAS meta-analysis of UK Biobank and Cohorts for Heart and Aging Research in Genomic Epidemiology cohorts (29,790 cases, 36,847 controls).^57^ Genotype-Tissue Expression (GTEx) v8 arterial eQTL data were obtained for aorta (n = 387), coronary artery (n = 213), and tibial artery (n = 584).^58^

### Stratified LD score regression

Cell-type marker genes were identified separately for each broad and fine cell type in scRNA-seq and snRNA-seq data. Expression specificity was calculated from cell-type-averaged, log-normalized expression as the proportion of each gene’s summed expression attributable to that cell type, and the top 10% of genes ranked by this specificity score were retained as the cell type’s marker set. Gene sets were converted to genomic annotations by extending gene coordinates by ±100 kb, and LD scores were computed for chromosomes 1–22 using LDSC (v1.0.1) with European reference data from the 1000 Genomes Project Phase 3. GWAS summary statistics were munged for LDSC and restricted to HapMap3 variants outside the major histocompatibility complex. Partitioned heritability was estimated for each combination of trait (CAD, LAS, PAD, CaP), modality (scRNA-seq, snRNA-seq), and cell type by jointly modeling the focal annotation with the 97 baseline annotations. *P* values were adjusted using the BH method.

### Gene-level prioritization using locus-to-gene mapping and colocalization

Candidate genes were prioritized using L2G scores from the Open Targets Platform GraphQL API v4.^59^ Disease-level queries were performed for CAD (MONDO_0005010), LAS (MONDO_0005490), and PAD (MONDO_0005386). Genes with L2G scores ≥0.5 were retained, and duplicate trait-gene associations were collapsed by retaining the maximum L2G score. Colocalization between GWAS summary statistics and GTEx arterial eQTLs (aorta, coronary artery, and tibial artery) was performed using coloc.abf.^60^ Variants shared between each GWAS and eQTL dataset were tested separately for each of the 12 trait-artery combinations using prior probabilities of p₁ = 10⁻⁴, p₂ = 10⁻⁴, and p₁₂ = 10⁻⁵. Genes with posterior probability for a shared causal variant (PP.H4) >0.8 were considered colocalized.

### Marker gene enrichment and disease-state expression of L2G and colocalized genes

L2G-prioritized genes and colocalized genes were tested separately for enrichment among cell type-specific marker genes defined for each broad and fine annotation. Background gene sets were restricted to genes expressed in the corresponding modality and broad cell type. Enrichment was assessed using one-sided Fisher’s exact tests, with odds ratios and 95% confidence intervals estimated from 2 × 2 contingency tables. P values were adjusted using the BH method within each trait for L2G analyses and within each trait-artery combination for colocalization analyses. Cell-type-specific differential expression between symptomatic and asymptomatic plaques was assessed as described above. P values were adjusted using BH method across pooled L2G-prioritized or colocalized genes, separately for each evidence class, cell type and modality.

### Mendelian randomization

Colocalized genes were further tested using two-sample cis-MR. Instruments were selected from GTEx arterial eQTLs at P<1×10⁻⁵ and LD-clumped using PLINK with the European 1000 Genomes reference panel at r²<0.1 within a 10,000 kb window. Exposure and outcome data were harmonized using TwoSampleMR::harmonise_data().^61^ The corresponding trait GWAS (CAD, LAS, PAD, CaP) served as outcome. Effect estimates were derived using the Wald ratio (single-instrument genes) or inverse-variance weighting (multi-instrument genes). P values were adjusted separately within each tissue-trait analysis using the BH method.

## Data Availability

All data produced in the present work are contained in the manuscript.

## Acknowledgments

The authors are grateful to all participants of the AtherOMICS Biobank for their participation in and contribution to this study. The authors also thank Jana Mattar, Stefan Mayrhofer, Paulo Alabarse, Mohamad Ali Antabi, Julian Louma, Panagiotis Zangas, Iulia Lupul, and Anastasia Osadcha, who contributed to the study operations of AtherOMICS.

## Supplementary figure legends

**Supplementary Fig. 1 Fine cell-state annotation in scRNA-seq and snRNA-seq carotid plaque datasets**.

UMAP embeddings of fine cell-state annotations in scRNA-seq and snRNA-seq datasets. Cells or nuclei are coloured by subtype-resolved annotation within smooth muscle, endothelial, macrophage, lymphoid and minor immune compartments. Fine annotations were assigned after broad lineage classification and within-lineage state resolution. UMAP, uniform manifold approximation and projection; scRNA-seq, single-cell RNA sequencing; snRNA-seq, single-nucleus RNA sequencing; SMC, smooth muscle cell; EC, endothelial cell; MP, macrophage; NK, natural killer; EndoMT, endothelial-to-mesenchymal transition; C1Q, complement component 1q

**Supplementary Fig. 2 Cell-type-specific expression concordance between scRNA-seq and snRNA-seq.**

Scatter plots showing mean normalized gene expression in scRNA-seq and snRNA-seq for shared genes within each broad cell type. Genes are coloured by relative enrichment in scRNA-seq or snRNA-seq, and selected modality-biased genes are labelled. Dotted lines indicate fold-change intervals around the dashed identity line. R² denotes the squared Pearson correlation coefficient for each cell type. scRNA-seq, single-cell RNA sequencing; snRNA-seq, single-nucleus RNA sequencing

**Supplementary Fig. 3 Pseudotime density distributions of symptomatic and asymptomatic plaques across smooth muscle cell, macrophage, T-cell and endothelial cell states.**

Ridgeline plots of normalized cell pseudotime density by fine cell-state, comparing symptomatic (red) and asymptomatic (blue) plaques, in scRNA-seq and snRNA-seq. Cell states are grouped by broad lineage (SMC, MP, T cell, EC). scRNA-seq, single-cell RNA sequencing; snRNA-seq, single-nucleus RNA sequencing; SMC, smooth muscle cell; MP, macrophage; EC, endothelial cell; EndoMT, endothelial-to-mesenchymal transition; Treg, regulatory T cell; C1Q, complement component 1q

**Supplementary Fig. 4| Cell-type enrichment and L2G-prioritised gene associations across CAD, LAS and PAD. a**, Stratified linkage disequilibrium score estimates of cell-type-specific heritability enrichment in scRNA-seq and snRNA-seq annotations. Point colour indicates heritability enrichment and point size indicates −log10-transformed FDR. **b**, Marker-gene enrichment of L2G-prioritised genes across cell types and traits. Point colour indicates log_2_ OR for enrichment and point size indicates −log10-transformed FDR. **c**, Differential expression of L2G-prioritised genes between symptomatic and asymptomatic plaque samples in scRNA-seq and snRNA-seq data. Colour indicates log_2_ fold change (Symptomatic − Asymptomatic); circles denote FDR < 0.05. The accompanying evidence matrix indicates whether each gene was detected in the corresponding trait-specific L2G set; filled circles indicate evidence present and open circles indicate evidence absent. scRNA-seq, single-cell RNA sequencing; snRNA-seq, single-nucleus RNA sequencing; SMC, smooth muscle cell; EC, endothelial cell; MP, macrophage; NK, natural killer; EndoMT, endothelial-to-mesenchymal transition; C1Q, complement component 1q; CAD, coronary artery disease; LAS, large artery stroke; PAD, peripheral artery disease; CaP, carotid plaque; FDR, false discovery rate; OR, odds ratio; L2G, locus-to-gene; FC, fold change; Sym, symptomatic; Asym, asymptomatic.

## REFERENCES

1. Global, regional, and national burden of stroke and its risk factors, 1990–2021: a systematic analysis for the Global Burden of Disease Study 2021. Lancet Neurol. 23, 973–1003 (2024).

2. Gleissner, C. A. Translational atherosclerosis research: From experimental models to coronary artery disease in humans. Atherosclerosis 248, 110–116 (2016).

3. Wolf, D. & Ley, K. Immunity and Inflammation in atherosclerosis. Circ. Res. 124, 315–327 (2019).

4. Virmani, R., Burke, A. P., Farb, A. & Kolodgie, F. D. Pathology of the vulnerable plaque. J. Am. Coll. Cardiol. 47, C13–18 (2006).

5. Kawai, K., Kawakami, R., Finn, A. V. & Virmani, R. Differences in Stable and Unstable Atherosclerotic Plaque. Arterioscler. Thromb. Vasc. Biol. 44, 1474–1484 (2024).

6. Depuydt, M. A. C. et al. Microanatomy of the Human Atherosclerotic Plaque by Single-Cell Transcriptomics. Circ. Res. 127, 1437–1455 (2020).

7. Pauli, J. et al. Single cell spatial transcriptomics integration deciphers the morphological heterogeneity of atherosclerotic carotid arteries. Nat. Commun. 16, 11282 (2025).

8. Fernandez, D. M. et al. Single-cell immune landscape of human atherosclerotic plaques. Nat. Med. 25, 1576–1588 (2019).

9. Wirka, R. C. et al. Atheroprotective roles of smooth muscle cell phenotypic modulation and the TCF21 disease gene as revealed by single-cell analysis. Nat. Med. 25, 1280–1289 (2019).

10. Pan, H. et al. Single-Cell Genomics Reveals a Novel Cell State During Smooth Muscle Cell Phenotypic Switching and Potential Therapeutic Targets for Atherosclerosis in Mouse and Human. Circulation 142, 2060–2075 (2020).

11. Dib, L. et al. Lipid-associated macrophages transition to an inflammatory state in human atherosclerosis increasing the risk of cerebrovascular complications. Nat. Cardiovasc. Res. 2, 656–672 (2023).

12. Živković, L., et al. Multimodal profiling of atherosclerosis: Protocol and pilot data for the AtherOMICS biobank. Sci. Adv. 12, eaee2639 (2026).

13. Traeuble, K. et al. Integrated single-cell atlas of human atherosclerotic plaques. Nat. Commun. 16, 8255 (2025).

14. Slysz, J., et al. Single-cell profiling reveals inflammatory polarization of human carotid versus femoral plaque leukocytes. JCI Insight 8, (2023).

15. Depuydt, M. A. C. et al. Single-cell T cell receptor sequencing of paired human atherosclerotic plaques and blood reveals autoimmune-like features of expanded effector T cells. Nat. Cardiovasc. Res. 2, 112–125 (2023).

16. Salido, E. et al. The 9p21.3 Coronary Artery Disease Risk Locus Drives Vascular Smooth Muscle Cells to an Osteochondrogenic State. Arterioscler. Thromb. Vasc. Biol. 45, 702–721 (2025).

17. Sanz, I. et al. Challenges and Opportunities for Consistent Classification of Human B Cell and Plasma Cell Populations. Front. Immunol. 10, (2019).

18. Domanska, D. et al. Single-cell transcriptomic analysis of human colonic macrophages reveals niche-specific subsets. J. Exp. Med. 219, e20211846 (2022).

19. Finucane, H. K. et al. Partitioning heritability by functional annotation using genome-wide association summary statistics. Nat. Genet. 47, 1228–1235 (2015).

20. Bashore, A. C. et al. High-Dimensional Single-Cell Multimodal Landscape of Human Carotid Atherosclerosis. Arterioscler. Thromb. Vasc. Biol. 44, 930–945 (2024).

21. Bashore, A. C., Coronel, J., Xue, C., Zhu, L. Y. & Reilly, M. P. Single-Cell Multimodal Profiling Reveals a Novel CD26+ Fibroblast Subpopulation in Atherosclerosis—Brief Report. Arterioscler. Thromb. Vasc. Biol. 45, 1389–1397 (2025).

22. Fidler, T. P. et al. Suppression of IL-1β promotes beneficial accumulation of fibroblast-like cells in atherosclerotic plaques in clonal hematopoiesis. Nat. Cardiovasc. Res. 3, 60–75 (2024).

23. Li, Y. et al. Down-regulated RGS5 by genetic variants impairs endothelial cell function and contributes to coronary artery disease. Cardiovasc. Res. 117, 240–255 (2021).

24. Tamiato, A. et al. Age-Dependent RGS5 Loss in Pericytes Induces Cardiac Dysfunction and Fibrosis. Circ. Res. 134, 1240–1255 (2024).

25. Cheng, W. et al. Regulator of G-protein signalling 5 protects against atherosclerosis in apolipoprotein E-deficient mice. Br. J. Pharmacol. 172, 5676–5689 (2015).

26. Carlström, M., Weitzberg, E. & Lundberg, J. O. Nitric Oxide Signaling and Regulation in the Cardiovascular System: Recent Advances. Pharmacol. Rev. 76, 1038–1062 (2024).

27. Kessler, T. et al. Functional characterization of the GUCY1A3 coronary artery disease risk locus. Circulation 136, 476–489 (2017).

28. Mauersberger, C. et al. Loss of soluble guanylyl cyclase in platelets contributes to atherosclerotic plaque formation and vascular inflammation. Nat. Cardiovasc. Res. 1, 1174–1186 (2022).

29. Kuzuya, M. et al. Effect of MMP-2 Deficiency on Atherosclerotic Lesion Formation in ApoE-Deficient Mice. Arterioscler. Thromb. Vasc. Biol. 26, 1120–1125 (2006).

30. Stöhr, R. et al. Loss of TIMP3 exacerbates atherosclerosis in ApoE null mice. Atherosclerosis 235, 438–443 (2014).

31. Mocci, G. et al. Single-Cell Gene-Regulatory Networks of Advanced Symptomatic Atherosclerosis. Circ. Res. 134, 1405–1423 (2024).

32. Turner, A. W. et al. Single-nucleus chromatin accessibility profiling highlights regulatory mechanisms of coronary artery disease risk. Nat. Genet. 54, 804–816 (2022).

33. Evrard, S. M. et al. Endothelial to mesenchymal transition is common in atherosclerotic lesions and is associated with plaque instability. Nat. Commun. 7, 11853 (2016).

34. Slenders, L. et al. Endothelial-to-mesenchymal transition gene signature derived from single-cell transcriptomics of human atherosclerotic tissue associates with stable plaque histological characteristics. Vascul. Pharmacol. 159, 107498 (2025).

35. Dy, D. C. M. et al. Twist1 Promotes Endothelial Phenotypic Transition and Unstable Plaque Phenotype During Atherosclerosis. Arterioscler. Thromb. Vasc. Biol. 46, e324428 (2026).

36. Hauseman, Z. J. et al. Structure of the MRAS–SHOC2–PP1C phosphatase complex. Nature 609, 416–423 (2022).

37. Bleckwehl, T. et al. Encompassing view of spatial and single-cell RNA sequencing renews the role of the microvasculature in human atherosclerosis. Nat. Cardiovasc. Res. 4, 26–44 (2025).

38. Chamberlin, J. T., Lee, Y., Marth, G. T. & Quinlan, A. R. Differences in molecular sampling and data processing explain variation among single-cell and single-nucleus RNA-seq experiments. Genome Res. 34, 179–188 (2024).

39. Denisenko, E. et al. Systematic assessment of tissue dissociation and storage biases in single-cell and single-nucleus RNA-seq workflows. Genome Biol. 21, 130 (2020).

40. Tritschler, S. et al. Concepts and limitations for learning developmental trajectories from single cell genomics. Development 146, dev170506 (2019).

41. Hafemeister, C. & Satija, R. Normalization and variance stabilization of single-cell RNA-seq data using regularized negative binomial regression. Genome Biol. 20, 296 (2019).

42. McGinnis, C. S., Murrow, L. M. & Gartner, Z. J. DoubletFinder: Doublet Detection in Single-Cell RNA Sequencing Data Using Artificial Nearest Neighbors. Cell Syst. 8, 329–337.e4 (2019).

43. Korsunsky, I. et al. Fast, sensitive and accurate integration of single-cell data with Harmony. Nat. Methods 16, 1289–1296 (2019).

44. Crow, M., Paul, A., Ballouz, S., Huang, Z. J. & Gillis, J. Characterizing the replicability of cell types defined by single cell RNA-sequencing data using MetaNeighbor. Nat. Commun. 9, 884 (2018).

45. Phipson, B. et al. propeller: testing for differences in cell type proportions in single cell data. Bioinformatics 38, 4720–4726 (2022).

46. Street, K. et al. Slingshot: cell lineage and pseudotime inference for single-cell transcriptomics. BMC Genomics 19, 477 (2018).

47. Roux de Bézieux, H., Van den Berge, K., Street, K. & Dudoit, S. Trajectory inference across multiple conditions with condiments. Nat. Commun. 15, 833 (2024).

48. Robinson, M. D., McCarthy, D. J. & Smyth, G. K. edgeR: a Bioconductor package for differential expression analysis of digital gene expression data. Bioinformatics 26, 139–140 (2010).

49. Liberzon, A. et al. The Molecular Signatures Database Hallmark Gene Set Collection. Cell Syst. 1, 417–425 (2015).

50. Korotkevich, G., Sukhov, V. & Sergushichev, A. Fast gene set enrichment analysis. 060012 Preprint at 10.1101/060012 (2019).

51. Law, C. W., Chen, Y., Shi, W. & Smyth, G. K. voom: precision weights unlock linear model analysis tools for RNA-seq read counts. Genome Biol. 15, R29 (2014).

52. Wu, D. & Smyth, G. K. Camera: a competitive gene set test accounting for inter-gene correlation. Nucleic Acids Res. 40, e133 (2012).

53. Bankhead, P. et al. QuPath: Open source software for digital pathology image analysis. Sci. Rep. 7, 16878 (2017).

54. Nelson, C. P. et al. Association analyses based on false discovery rate implicate new loci for coronary artery disease. Nat. Genet. 49, 1385–1391 (2017).

55. Mishra, A. et al. Stroke genetics informs drug discovery and risk prediction across ancestries. Nature 611, 115–123 (2022).

56. Klarin, D. et al. Genome-wide association study of peripheral artery disease in the Million Veteran Program. Nat. Med. 25, 1274–1279 (2019).

57. Omarov, M., et al. Automated Deep Learning-Based Detection of Early Atherosclerotic Plaques in Carotid Ultrasound Imaging. Preprint at 10.1101/2024.10.17.24315675 (2025).

58. THE GTEX CONSORTIUM. The GTEx Consortium atlas of genetic regulatory effects across human tissues. Science 369, 1318–1330 (2020).

59. Ghoussaini, M. et al. Open Targets Genetics: systematic identification of trait-associated genes using large-scale genetics and functional genomics. Nucleic Acids Res. 49, D1311–D1320 (2021).

60. Giambartolomei, C. et al. Bayesian Test for Colocalisation between Pairs of Genetic Association Studies Using Summary Statistics. PLOS Genet. 10, e1004383 (2014).

61. Hemani, G. et al. The MR-Base platform supports systematic causal inference across the human phenome. eLife 7, e34408 (2018).

