## Supplementary figures and images for "Single-cell profiling of human plaques reveals cellular and molecular signatures of symptomatic atherosclerosis"

### Supplemental Figures

#### Slide 1
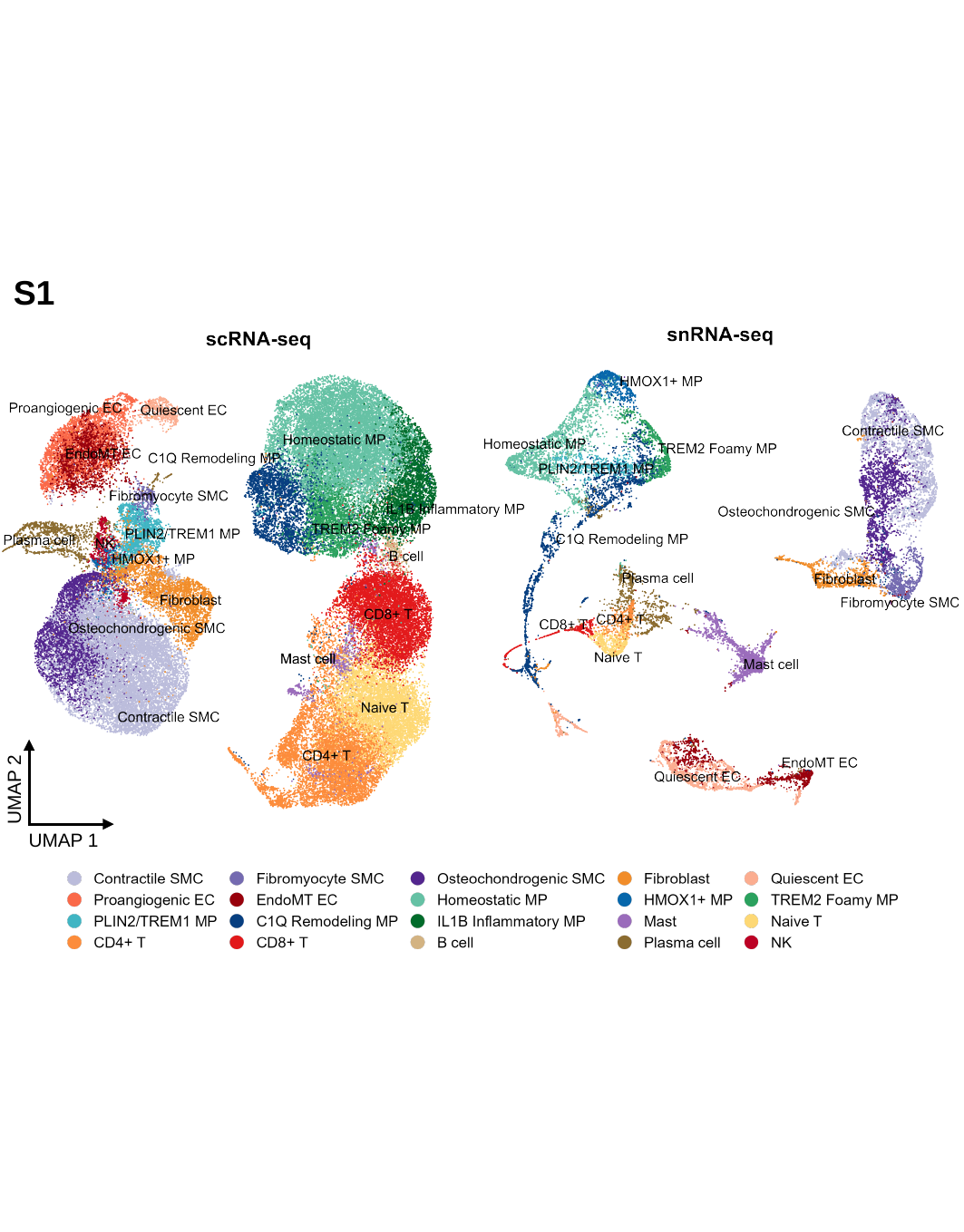

S1
UMAP 2
UMAP 1

#### Slide 2
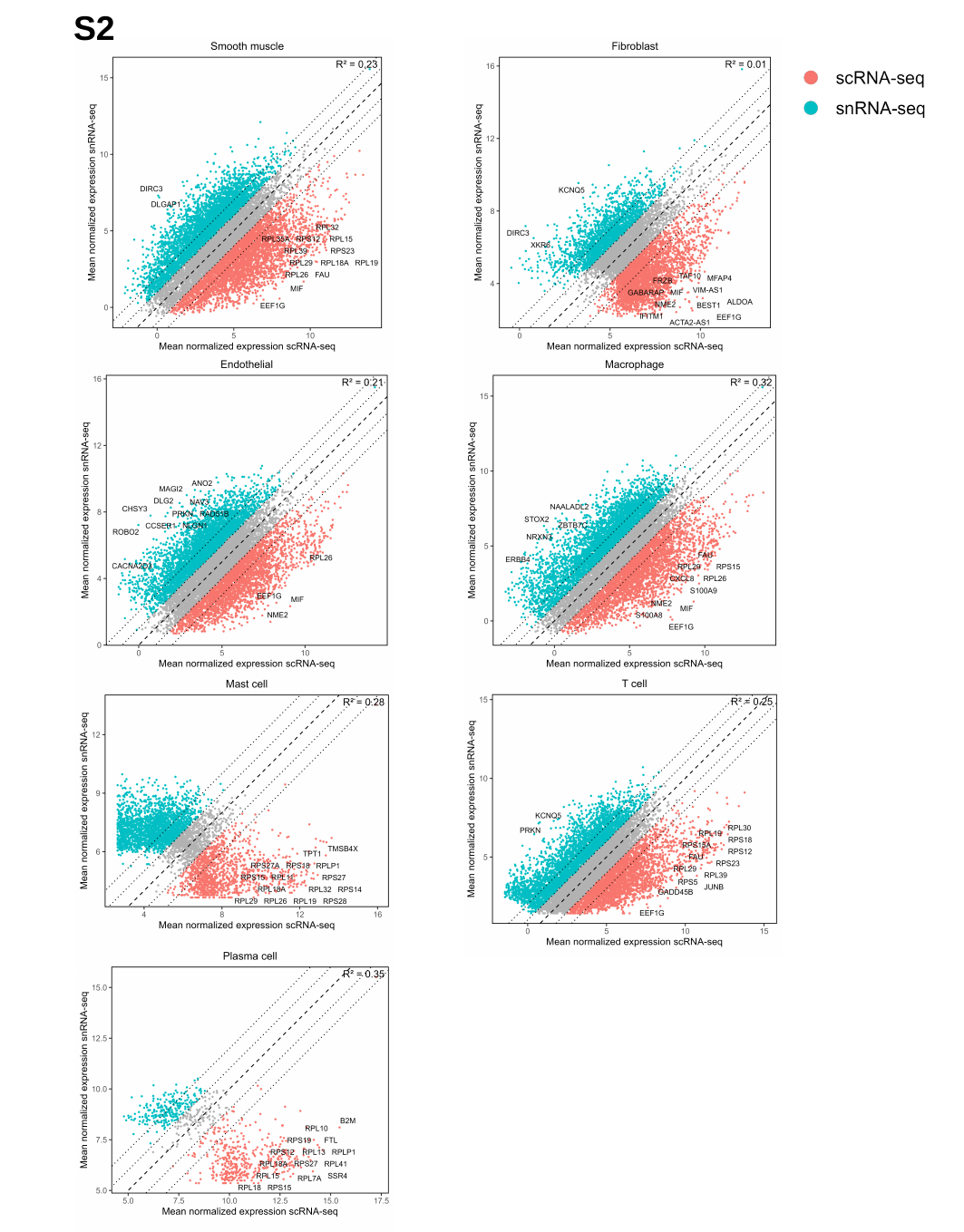

S2

#### Slide 3
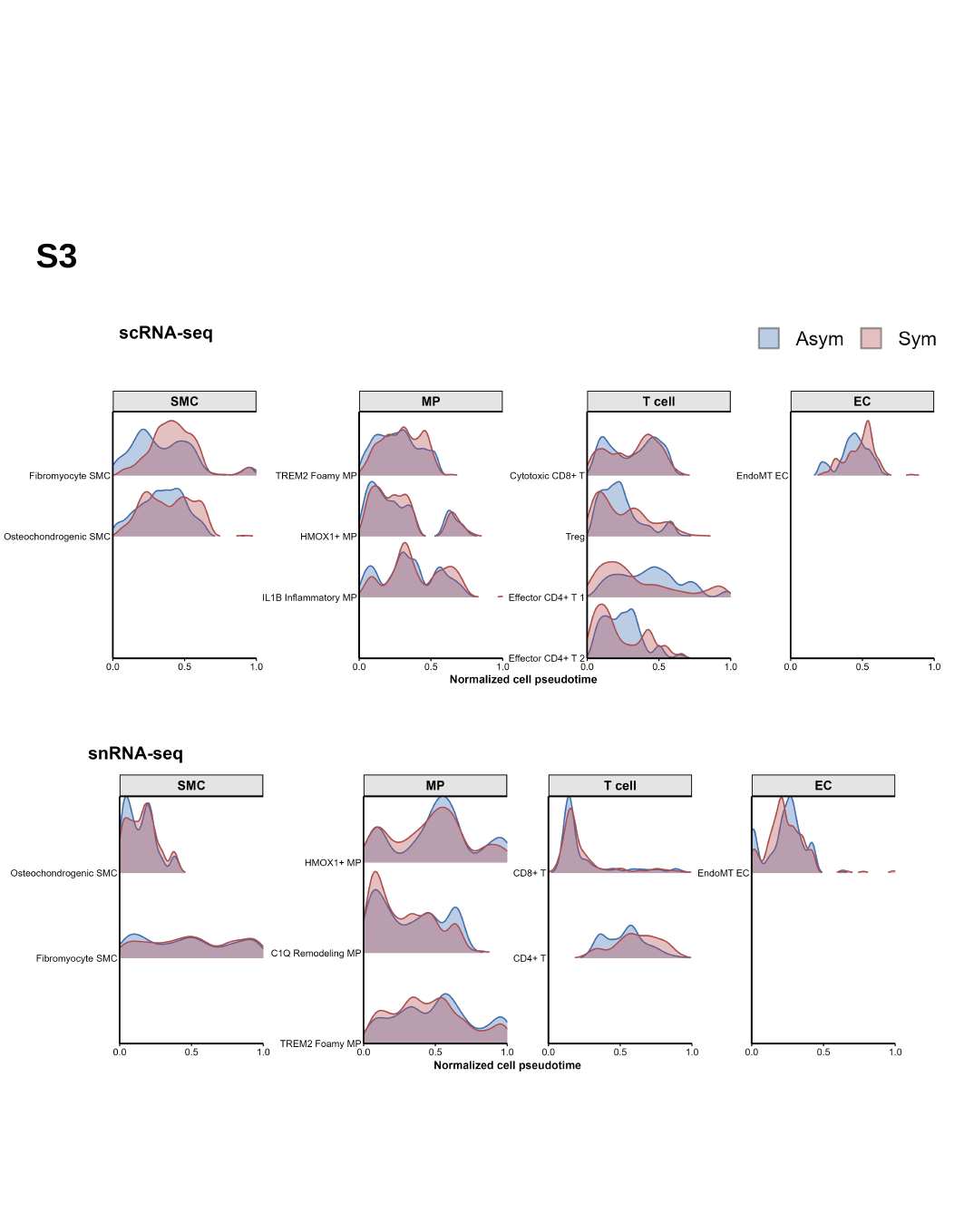

S3

#### Slide 4
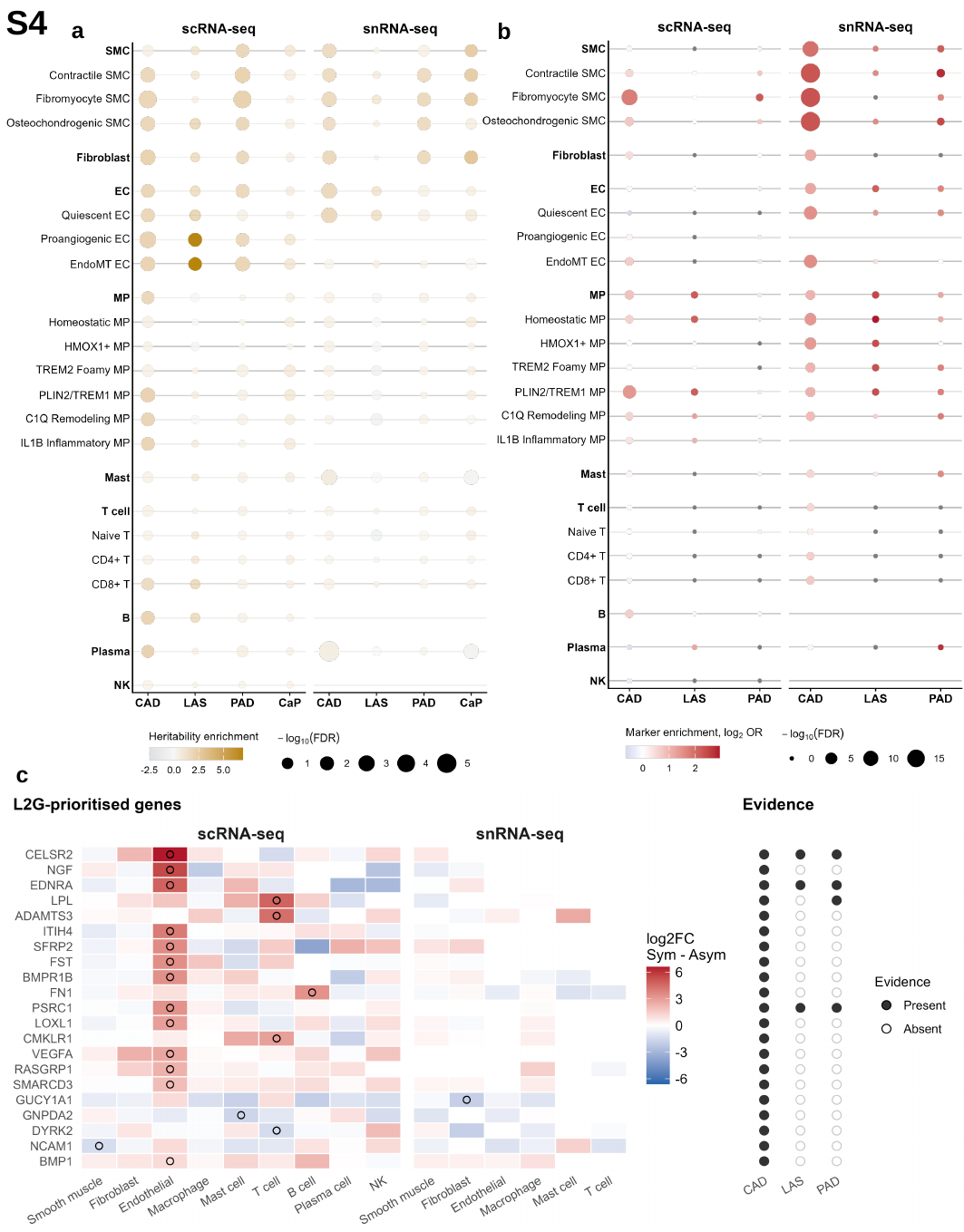

S4
a
b
c
